# Engagement with the HCV care cascade among high-risk groups: a population-based study

**DOI:** 10.1101/2022.10.29.22281692

**Authors:** Aysegul Erman, Karl Everett, William W. L. Wong, Farinaz Forouzannia, Christina Greenaway, Naveed Janjua, Jeffrey C. Kwong, Beate Sander

**Affiliations:** Toronto Health Economics and Technology Assessment Collaborative (THETA), University Health Network, Toronto, ON, Canada; ICES, Toronto, Ontario, Canada; School of Pharmacy, University of Waterloo, Kitchener, ON, Canada; Division of Infectious Diseases, Jewish General Hospital, McGill University, Montreal, QC, Canada; British Columbia Centre for Disease Control (BCDC), Vancouver, BC, Canada; University of Toronto; Public Health Ontario, Toronto, Ontario, Canada

**Keywords:** health services research, chronic hepatitis C, immigrants, residential instability, baby-boomer, care cascade, administrative data, population-based study, real-world data, cause-specific hazard

## Abstract

**Background:** Hepatitis C virus (HCV) elimination requires a thorough understanding of the care cascade. A direct-acting-antiviral (DAA)-era description of the care cascade has not been undertaken in Ontario, Canada. Our primary objective was to describe the current population-level care cascade in the general Ontario population and among key risk-groups — baby-boomers, immigrants, and individuals experiencing residential instability. The secondary objective was to identify predictors of engagement.

**Methods:** We conducted a population-based cohort study of Ontario residents undergoing HCV testing between January 1, 1999, and December 31, 2018, and mapped the care cascade [antibody diagnosed, RNA tested, RNA positive, genotyped, treated, achieved sustained virologic response (SVR), reinfected/relapsed] as of December 31, 2018. The cascade was stratified by risk groups. Cause-specific hazard modeling was used to identify demographic, and socioeconomic predictors of engagement with key steps of the cascade.

**Results:** Among 108,428 Ontario resident living with an HCV antibody diagnosis, 88% received confirmatory RNA testing; of these, 62% tested positive and 94% of positive tests were genotyped. Of those with confirmed viremia, 53% initiated treatment and 76% of treated individuals achieved SVR, while ∼1% experienced reinfection or relapse. Males, older birth cohorts, long-term residents, those with a history of substance use disorder and social marginalisation (e.g., material deprivation, residential instability), and those initially diagnosed in the pre-DAA era exhibited lower rates of engagement with almost every step of HCV care.

**Conclusions:** Despite DAA-era improvements, treatment initiation remains a major gap. HCV screening and linkage-to-treatment, particularly for those with a history of substance use disorder and social marginalisation, will be needed to equitably close gaps in HCV care in Ontario.

## Introduction

Introduction of highly effective direct-acting antivirals (DAAs) has revolutionized the treatment of Hepatitis C virus (HCV). These drugs offer an opportunity to eliminate HCV as a public health concern. Thus, the World Health Organization (WHO) has set ambitious global elimination targets (e.g., 90% reduction in incidence and 65% in mortality by 2030) (1).

Meeting these targets will require a thorough understanding of the gaps along the “HCV care cascade” from diagnosis to the achievement of sustained virologic response (SVR). However, individuals living with hepatitis C are highly heterogeneous. For instance, key risk groups in Canada include foreign-born individuals, baby-boomers (i.e., individuals born between 1945 and 1965), people who inject drugs, people who are homeless, incarcerated individuals, men who have sex with men, and First Nations groups, each have unique challenges in accessing and remaining engaged in healthcare (2–6). In addition to scaling up the HCV care cascade, elevated risks of reinfection for certain marginalised populations means that engagement with HCV care beyond SVR achievement may be warranted (7–9).

In Ontario, Canada’s most populous province, the availability of large population-based datasets allows for the creation of real-world care cascades that provide a more complete picture of the state of HCV care. In deed, previous work using Ontario health administrative data has mapped the pre-DAA era HCV care cascade by immigration status between January 1999 and December 2014 (2). However, the care cascade in the DAA era (i.e., beyond 2014) has not been described, for either the general population or for key risk groups.

The primary objective of this paper was to describe the real-world HCV care cascade in Ontario, particularly focusing on three risk groups 1 baby-boomers, immigrants, and individuals experiencing residential instability. The secondary objective was to identify factors associated with engagement with key steps of the care cascade.

## Methods

### Study population and setting

In Ontario, Canada, confirmation of reportable diseases typically occurs at Public Health Ontario (PHO) laboratories where the vast majority (95%) of HCV testing and all nucleic acid testing for viral hepatitis is processed. We included all Ontario residents who underwent testing for HCV antibody and RNA at Public Health Ontario (PHO) laboratories between January 1, 1999, and December 31, 2018. In total, we linked the HCV laboratory testing records of 1,597,571 individuals with health administrative and demographic data held at ICES and identified 134,655 individuals with laboratory data consistent with HCV antibody positivity. For our primary objective, we mapped the HCV care cascade for 108,428 individuals who were alive as of December 31, 2018, to obtain a snapshot of the most current state of HCV care in the province (following the exclusion of 26,227 individuals who died on or before 2018, consistent with previous cascade studies(2,10)). For our secondary objective, we included all 134,655 individuals engaged with HCV care during the study period to identify factors associated with engagement.

### Data sources

Hepatitis testing data including HCV antibody testing, HCV RNA testing, genotyping, and hepatitis B virus (HBV) testing status were collected from the PHO laboratory dataset. Data on HCV antiviral dispensation were collected from the Ontario Drug Benefit (ODB) database using drug identification numbers (DIN) listed in **Supplemental Table 1**. Immigration status was collected from the Immigration, Refugees, and Citizenship Canada (IRCC) permanent Resident Database, which holds records of individuals who have been granted permanent resident status in Canada since 1985. Demographic information on birth year, gender, age, rurality, and neighborhood income quintile was obtained from the Registered Persons Database (RPDB). Markers of social marginalisation such as residential instability quintile, material deprivation quintile, and ethnic concentration quintile were obtained from the Ontario Marginalisation Index Database (ONMARG). HBV infection status was determined based on hepatitis B surface antigen (HBsAg) test results. Information on HIV status, cirrhosis, decompensated cirrhosis (DC), hepatocellular carcinoma (HCC), liver transplant, death, substance use disorders related to drug and/or alcohol use and comorbidities (Aggregated Diagnosis Groups [ADG] comorbidity classification scheme) were collected from the National Ambulatory Care Reporting System (NACRS), Ontario Health Insurance Program (OHIP), Canadian Institute for Health Information Discharge Abstract Database (DAD), Ontario Mental Health Reporting System (OMHRS), Ontario Cancer Registry (OCR), and Office of the Registrar General Death registry (ORGD) datasets, using diagnostic, procedure-related and death codes listed in **Supplemental Table 2A-2B**. Baseline characteristics were collected on index date of first engagement with the HCV care cascade and at the time of treatment initiation. The Johns Hopkins Adjusted Clinical Group® (ACG®) system was used to determine the total number ADGs over the one-year period preceding index (11). All records were linked using unique identifiers and analysed at ICES.

### HCV care cascade

The HCV care cascade stages are described in **Table 1**. We mapped care cascades for three key risk groups 1) immigrants, 2) baby-boomers, and 3) individuals experiencing residential instability. Immigrants were defined as those who received Canadian permanent resident status after January 1985. Baby-boomers were defined as individuals born between 1945 and 1965. Individuals falling outside of this birth cohort were divided into those born before 1945 and those born after 1965. Residential instability (i.e., the level of family and housing instability) was defined by the neighborhood-level residential instability quintile, grouped into low (quintiles 1 and 2), medium (quintile 3), and high (quintiles 4 and 5).

**Table 1.** HCV Care cascade.

|  |  |
| --- | --- |
| **[STAGE 1]**<br>**HCV antibody positive** | Number of individuals with at least one positive HCV AB test result, a record of HCV RNA testing, an HCV genotype or a record, or HCV antiviral drug dispensation. |
| **[STAGE 2]**<br>**HCV RNA tested** | Number of individuals with a record of HCV RNA testing, a record of HCV genotype, or a record of HCV antiviral drug dispensation. |
| **[STAGE 3]**<br>**HCV RNA positive** | Number of individuals with either a record of positive HCV RNA testing, a record of HCV genotype or a record of HCV antiviral drug dispensation. |
| **[STAGE 4]**<br>**HCV genotype tested** | Number of individuals with a record of HCV genotype or a record of HCV antiviral drug dispensation. |
| **[STAGE 5]**<br>**HCV antiviral treatment initiated** | Number of individuals with either a record of HCV antiviral drug dispensation from public drug plan or those presumably treated through private insurance or via out-of-pocket expenditures i.e., those without a record of HCV antiviral dispensation from public drug records but who have a final negative HCV RNA test result consistent with SVR achievement. |
| **[STAGE 6]**<br>**SVR achieved** | Number of individuals with a record of HCV antiviral drug dispensation from public drug plan who have a negative HCV RNA test result at least 12 weeks after the dispensation of the last HCV antiviral or those who presumably achieved SVR through private insurance i.e., those without a public record of HCV antiviral drug dispensation but who had a final negative HCV RNA test result consistent with SVR. |
| **[STAGE 7]**<br>**Reinfection or relapse on treatment** | Number of individuals with a record of HCV antiviral drug dispensation who have had at least one negative HCV RNA test result after the last antiviral dispensation on record which was followed by a final positive HCV RNA test result. |
| **[STAGE 8]**<br>**Acute resolved** | Number of individuals with a negative HCV RNA test result within 12 months following the first positive RNA test, and with no positive RNA test result within the first 12 months and no record of HCV antiviral use. |

### Statistical Analyses

To identify factors associated with time-to-engagement with key steps of the care cascade and to account for right censoring and variability of follow-up times, we used cause-specific hazard models to estimate covariate-adjusted hazard ratios (aHR) accounting for death as a competing risk [8]. Covariates were collected prior to event time. For the cause-specific hazard models, the individuals were followed from index date up to either the target event of interest (e.g., RNA testing, HCV treatment initiation, SVR), or until censoring at death or at the end of follow-up (31^st^ December 2018). Time origin was chosen as the first date on record of the previous cascade step. For instance, time-to-RNA-testing was estimated from the first date of HCV antibody testing; time-to-treatment-initiation from the first date of HCV RNA positivity, time-to-SVR was estimated from the first date of HCV treatment initiation (i.e., antiviral dispensation)[9]. For individuals presumed to have initiated treatment through private means (i.e. those with missing data on the drug dispensation from public drug plan records), date of treatment initiation was imputed based on the date of last HCV RNA positive test on record. As a supplementary analysis, logistic regression was performed to identify predictors of HCV antibody positivity and RNA test positivity in the province using data on all individuals who received HCV antibody (N=1,597,571) or HCV RNA testing (N=113,422), respectively. Predictors included in the logistic and hazard models included baseline age at engagement with the cascade, sex, rurality, birth year, comorbidity profile, neighborhood income, immigrant status, residential instability level, neighborhood income level, material deprivation level, ethnic concentration level, DAA-era (vs. pre-DAA) engagement with the care cascade, HIV or HBV coinfection, substance use disorder, and liver disease severity at baseline. SVR was also adjusted for having ever been retreated with HCV antivirals. Covariates were selected *a priori based* on previous studies(2,14). All statistical analyses were performed using SAS Enterprise Guide 7.15 (SAS Institute, Inc.).

## Results

### Study cohort

The HCV care cascade included a total of 108,428 individuals who were either living with or had recovered from HCV infection during the study time frame. Forty-nine percent belonged to the baby-boomer generation, 46% were born after 1965 and 5% were born before 1945. The majority were long-term residents of Canada (85%), and the remainder immigrated to Canada after 1985. More than half (57%) resided in neighborhoods with the highest levels of residential instability (**Table 2**).

**Table 2.**
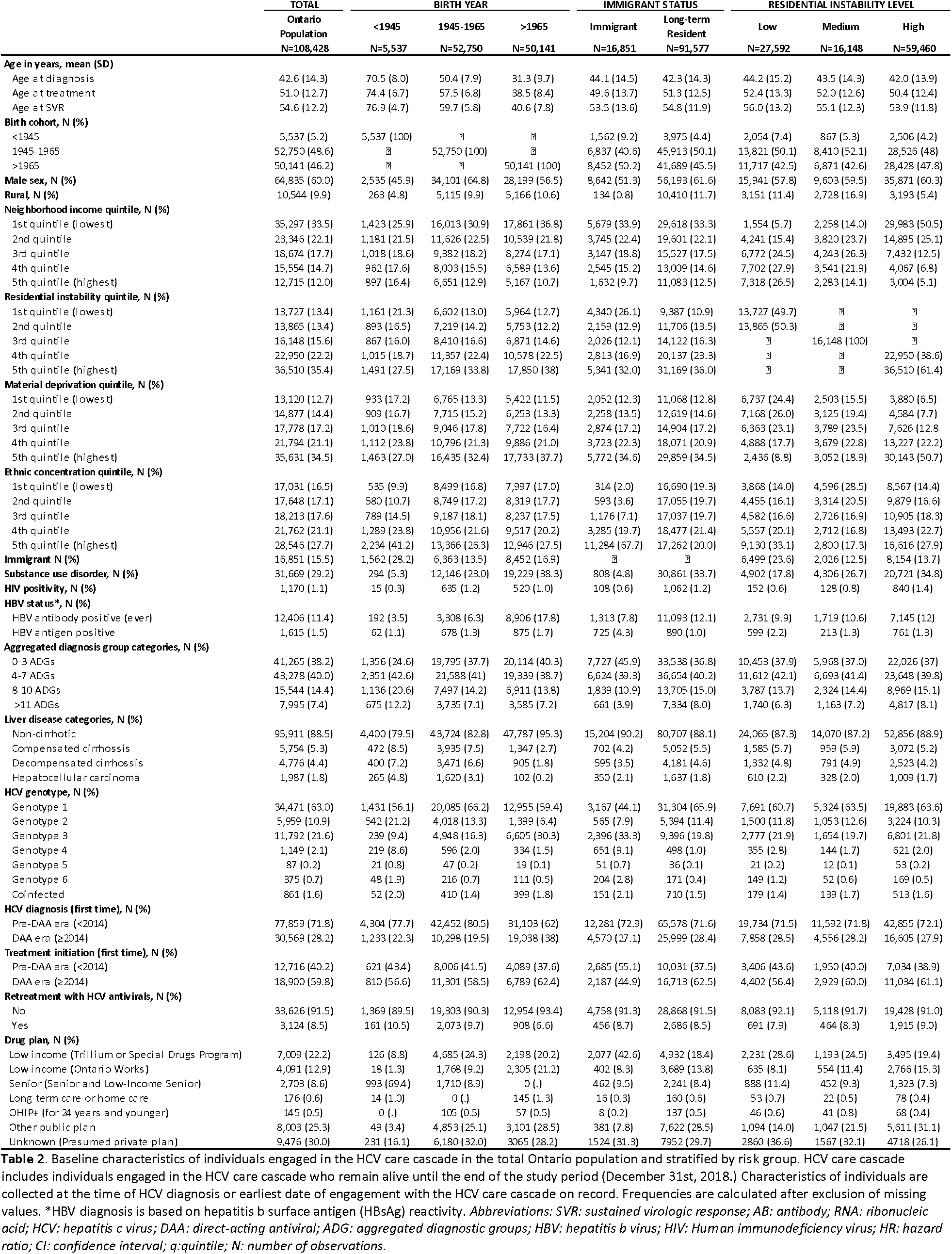
Characteristics of individuals engaged in HCV care cascade stratified by risk groups.

On average, Ontario residents were aged 43 years at initial HCV diagnosis, 51 years at the time of treatment initiation, and 55 years at SVR. The majority were male (60%), resided in an urban setting (>90%), were in the lowest two income quintiles (55%), and the top two material deprivation quintiles (55%). Twenty-nine percent had a history of substance use disorder related to drug and/or alcohol use, 1.1% were coinfected with HIV and 1.5% with HBV at the time of diagnosis. At the time of HCV diagnosis, 5.3% had been diagnosed with compensated cirrhosis, 4.4% with DC, and 1.8% with HCC. Genotype 1 was the most common viral strain (63%).

Although most of the cohort was initially diagnosed in the pre-DAA era (72%), HCV treatment was most frequently initiated for the first time during the DAA era (60%). Overall, 38% of those who initiated treatment had a history of drug- and/or alcohol-related substance use disorder, 2% were HIV-coinfected, 24% had compensated cirrhosis, and ∼8% had advanced liver disease (DC or HCC) at the time of treatment (**S.Table 3**). For those initiating treatment through Ontario’s public drug plan, 50% accessed antivirals through programs for those in financial need and/or those facing drug costs disproportionate to income (e.g., Trillium or Special Drugs Program, Ontario Works), corresponding to 35% of all treated individuals. Individuals experiencing reinfection or relapse tended to be younger at treatment compared to those who achieved SVR, and were also more likely to face social marginalisation, substance use disorder, HIV coinfection, and were more likely to have had advanced liver disease at the time of treatment (**S.Table 3**). Demographic characteristics by care cascade engagement for all risk groups are displayed in **S.Tables 4**-**6**.

### HCV care cascades

The HCV care cascade for the study cohort is displayed in **Figure 1A**. We identified 108,428 individuals alive and with a positive HCV antibody diagnosis as of 2018 in the province. Of antibody-diagnosed individuals, 95,002 (88%) were tested for RNA. Of those receiving RNA testing, 59,370 (62%) were RNA positive and of those testing positive 56,140 (94%) were genotyped. Among individuals testing RNA positive, 31,656 (53%) initiated treatment and 23,950 (76%) of those receiving treatment achieved SVR while 1% were reinfected or relapsed.

**Figure 1.**
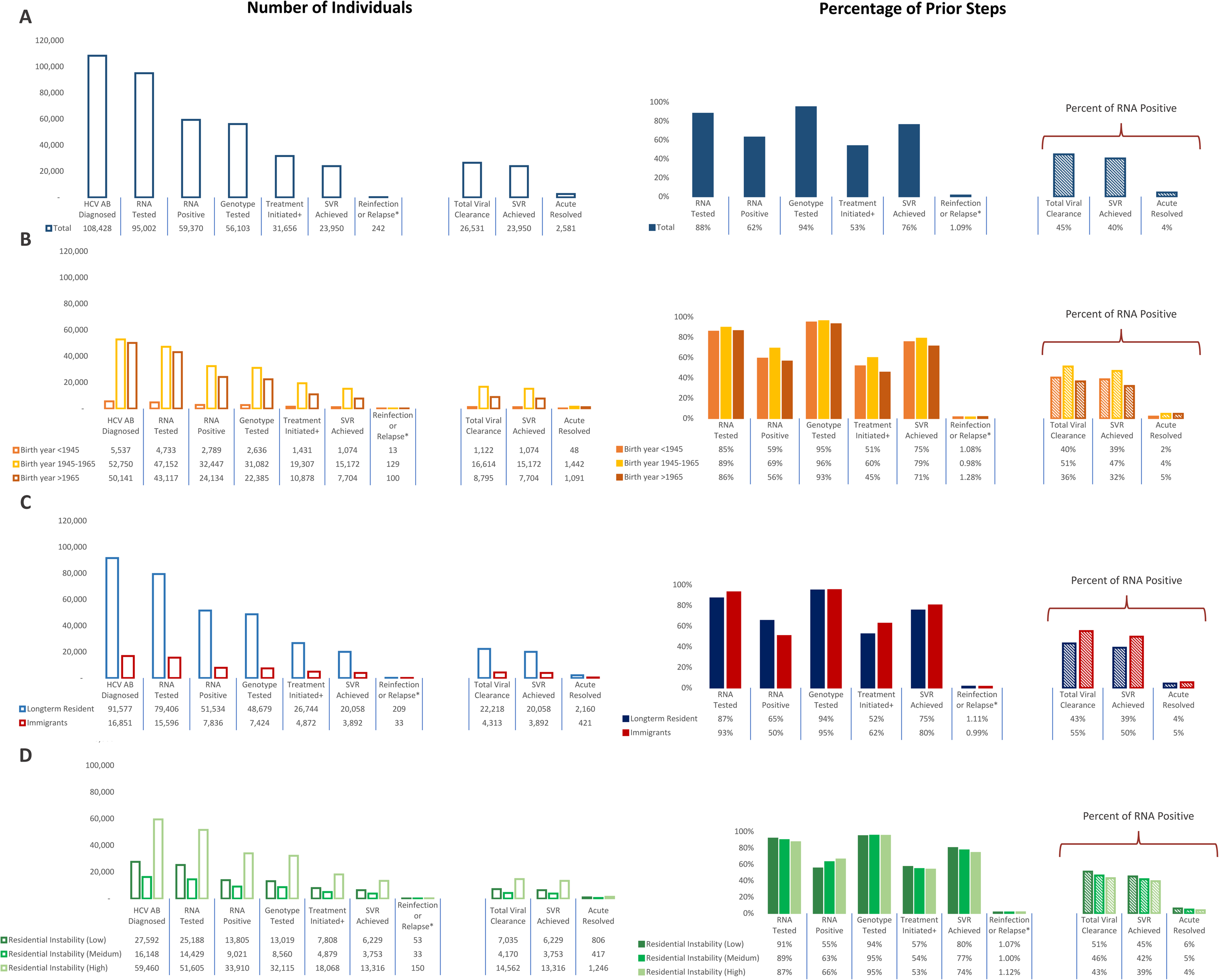
The level of HCV care cascade engagement among key risk groups. Figure showing the population level HCV care cascade in Ontario (A) for all individuals ever diagnosed with hepatitis C from January 1999 onwards and alive up to December 2018 inclusive and stratified by (B) birth year, (C) immigrant status, and (D) residential instability. *Percentage reinfection or relapse is presented as percent of all who initiated HCV antiviral treatment. Percent treatment initiation is presented as percentage of all RNA positive cases. *Abbreviations: SVR: sustained virologic response; AB: antibody; RNA: ribonucleic acid; HCV: hepatitis c virus*.

Generally, HCV confirmatory RNA testing level was not substantially different across the birth years (**Figure 1B)**. RNA positivity was also greater among the baby-boomers relative to the other birth cohorts as were treatment initiation and SVR, while the youngest birth cohort had the lowest level of treatment and SVR. Reinfection/relapse was also higher among the youngest birth cohort.

In general, immigrants tended to have slightly higher levels of confirmatory RNA testing and lower RNA test positivity compared to long-term residents (**Figure 1C)**. In addition, immigrants also had higher levels of treatment initiation and SVR. Finally, when we stratified the care cascade by residential instability (**Figure 1D**), we found decreasing levels of confirmatory RNA testing and increasing rates of RNA test positivity with increasing housing instability. Similarly, treatment initiation and SVR were also lower for those facing increasing levels of residential instability whereas reinfection/relapse was higher.

**Figure 2.**
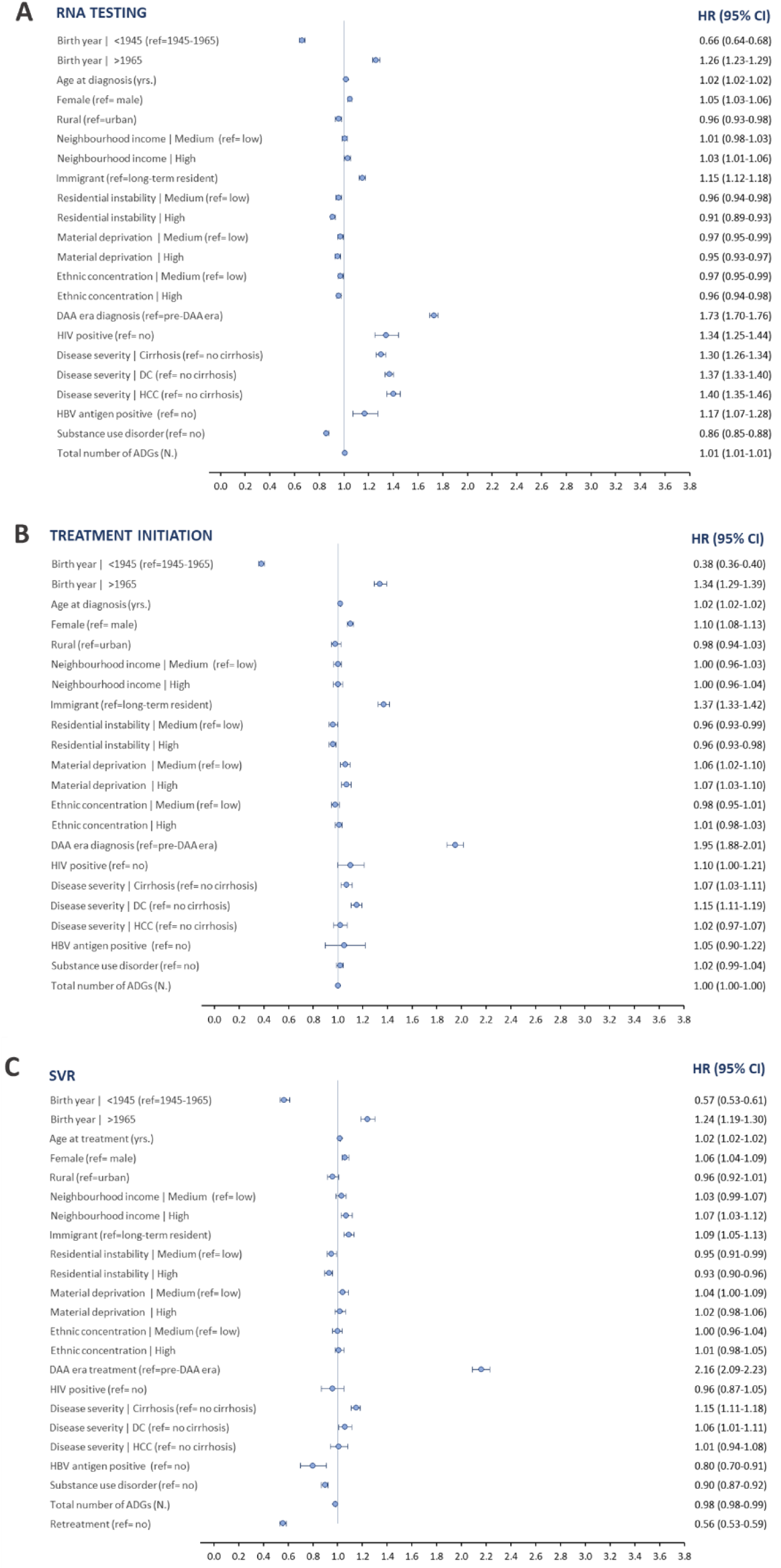
Covariate -adjusted hazard ratios for engagement with key stages of the HCV care cascade. Figure showing the predictors associates with (A) time to HCV RNA testing among HCV antibody positive cases, (B) time to treatment initiation among HCV RNA positive cases, (C) time to SVR among HCV treated cases. Cause-specific hazard models for time-to-event outcomes were used to estimate hazard ratios adjusting for multiple predictors. *Abbreviations: SVR: sustained virologic response; AB: antibody; RNA: ribonucleic acid; HCV: hepatitis c virus; DAA: direct-acting antiviral; ADG: aggregated* diagnostic groups; DC: decompensated cirrhosis; HCC: hepatocellular carcinoma; HBV: hepatitis b virus; HIV: Human immunodeficiency virus; HR: hazard ratio; CI: confidence interval.

### Odds of HCV antibody and RNA test positivity

In logistic regression analyses adjusting for multiple covariates, the odds of HCV antibody test positivity among those receiving antibody testing was significantly higher for those with a history of drug and alcohol-related substance use (aOR: 3.58, 95%CI, 3.52-3.63), and those with higher levels of residential instability (aOR: 1.16, 95%CI, 1.14-1.18) and material deprivation (aOR: 1.29, 95%CI, 1.27-1.32) (**Table 3**). The odds of antibody positivity was also higher for those presenting with cirrhosis (aOR:3.84, 95%CI, 3.71-3.96), DC (aOR: 2.35, 95%CI, 2.29-2.42), or HCC (aOR: 6.82, 95%CI, 6.52-7.15) compared to individuals without cirrhosis at the time of HCV diagnosis. Similarly, baby-boomers also displayed a higher odds of antibody positivity relative to other birth cohorts. Finally, HIV-coinfection was also associated with a higher odds of antibody positivity (aOR: 2.29, 95%CI, 2.12-4.24). In contrast, the odds of antibody test positivity were lower for females (aOR: 0.68, 95%CI, 0.67-0.68 vs. male), rural populations (aOR: 0.63, 95%CI, 0.61-0.64 vs. urban), those with highest income levels (aOR: 0.70, 95%CI, 0.69-0.72), immigrants (aOR: 0.91, 95%CI, 0.92-0.96 vs. long-term resident), individuals coinfected with HBV (aOR: 0.64, 95%CI, 0.59-0.68), and those diagnosed in the DAA era (aOR: 0.37, 95%CI, 0.27-0.38 vs. pre-DAA).The results of RNA positivity were consistent with those of antibody positivity, with the exception of HIV-coinfection, which was associated with a higher odds of antibody positivity, but a lower odds of RNA positivity (aOR: 0.67, 95%CI, 0.60-0.75).

**Table 3.** Predictors of HCV antibody and RNA test positivity.

| Covariates | AB Positivity<br>N=1,552,181 |  | RNA Positivity<br>N=108,952 |  |
| --- | --- | --- | --- | --- |
|  | Unadjusted<br>OR (95% CI) | Adjusted<br>OR (95% CI) | Unadjusted<br>OR (95% CI) | Adjusted<br>OR (95% CI) |
| **Age (yrs.)** |  |  |  |  |
| At time of testing | **1.01 (1.01-1.01)** | **0.99 (0.99-0.99)** | 1.00 (1.00-1.00) | **0.99 (0.99-0.99)** |
| **Birth year** |  |  |  |  |
| 1945-1965 [ref.] | ? | ? | ? | ? |
| <1945 | **0.36 (0.36-0.37)** | **0.44 (0.43-0.45)** | **0.72 (0.68-0.75)** | **1.13 (1.06-1.20)** |
| >1965 | **0.28 (0.28-0.28)** | **0.25 (0.25-0.26)** | **0.54 (0.53-0.55)** | **0.52 (0.49-0.54)** |
| **Sex** |  |  |  |  |
| Male [ref.] | ? | ? | ? | ? |
| Female | **0.52 (0.52-0.53)** | **0.68 (0.67-0.68)** | **0.60 (0.59-0.62)** | **0.69 (0.67-0.71)** |
| **Rurality** |  |  |  |  |
| Urban [ref.] | ? | ? | ? | ? |
| Rural | **0.79 (0.78-0.81)** | **0.63 (0.61-0.64)** | **1.18 (1.13-1.23)** | 1.02 (0.97-1.08) |
| **Neighborhood income quintile** |  |  |  |  |
| Low (q. 1-2) [ref.] | ? | ? | ? | ? |
| Medium (q. 3) | **0.70 (0.69-0.71)** | **0.85 (0.83-0.87)** | **0.83 (0.80-0.86)** | **0.90 (0.86-0.94)** |
| High (q. 4-5) | **0.52 (0.51-0.53)** | **0.70 (0.69-0.72)** | **0.70 (0.68-0.72)** | **0.77 (0.74-0.81)** |
| **Residential instability quintile** |  |  |  |  |
| Low (q. 1-2) [ref.] | ? | ? | ? | ? |
| Medium (q. 3) | **1.23 (1.21-1.26)** | **1.03 (1.01-1.05)** | **1.37 (1.32-1.43)** | **1.13 (1.08-1.18)** |
| High (q. 4-5) | **1.72 (1.70-1.75)** | **1.16 (1.14-1.18)** | **1.56 (1.51-1.60)** | **1.22 (1.18-1.26)** |
| **Material deprivation quintile** |  |  |  |  |
| Low (q. 1-2) [ref.] | ? | ? | ? | ? |
| Medium (q. 3) | **1.37 (1.34-1.39)** | **1.13 (1.11-1.15)** | **1.19 (1.15-1.24)** | **1.04 (1.00-1.09)** |
| High (q. 4-5) | **1.98 (1.96-2.01)** | **1.29 (1.27-1.32)** | **1.41 (1.37-1.45)** | **1.08 (1.04-1.13)** |
| **Ethnic concentration quintile** |  |  |  |  |
| Low (q. 1-2) [ref.] | ? | ? | ? | ? |
| Medium (q. 3) | **0.96 (0.94-0.97)** | **0.98 (0.96-1.00)** | **0.90 (0.86-0.93)** | **0.92 (0.88-0.96)** |
| High (q. 4-5) | **0.93 (0.91-0.94)** | **0.97 (0.96-0.99)** | **0.68 (0.66-0.70)** | **0.74 (0.72-0.77)** |
| **Immigrant status** |  |  |  |  |
| Long-term resident [ref.] | ? | ? | ? | ? |
| Immigrant | **0.69 (0.67-0.70)** | **0.94 (0.92-0.96)** | **0.53 (0.52-0.55)** | **0.82 (0.79-0.85)** |
| **Substance use disorder\*** |  |  |  |  |
| No [ref.] | ? | ? | ? | ? |
| Yes | **4.30 (4.25-4.36)** | **3.58 (3.52-3.63)** | **1.87 (1.82-1.92)** | **2.14 (2.07-2.22)** |
| **HIV positivity\*** |  |  |  |  |
| No [ref.] | ? | ? | ? | ? |
| Yes | **5.24 (4.91-5.60)** | **2.29 (2.12-2.47)** | **0.78 (0.70-0.87)** | **0.67 (0.60-0.75)** |
| **HBsAG positivity\*** |  |  |  |  |
| No [ref.] | ? | ? | ? | ? |
| Yes | **0.89 (0.84-0.95)** | **0.64 (0.59-0.68)** | **0.18 (0.16-0.20)** | **0.19 (0.16-0.21)** |
| **Total Number of ADGs (N.)\*** | **1.03 (1.02-1.03)** | **0.96 (0.95-0.96)** | **0.98 (0.98-0.99)** | **0.95 (0.95-0.95)** |
| **Liver disease\*** |  |  |  |  |
| No cirrhosis [ref.] | ? | ? | ? | ? |
| Compensated cirrhosis | **5.87 (5.71-6.04)** | **3.84 (3.71-3.96)** | **2.10 (1.98-2.23)** | **2.08 (1.95-2.22)** |
| Decompensated cirrhosis | **4.92 (4.81-5.04)** | **2.35 (2.29-2.42)** | **2.28 (2.17-2.40)** | **1.98 (1.87-2.09)** |
| Hepatocellular carcinoma | **9.92 (9.53-10.3)** | **6.82 (6.52-7.15)** | **3.30 (3.02-3.60)** | **3.08 (2.80-3.38)** |
| **HCV testing (first time)** |  |  |  |  |
| Pre-DAA era (<2014) [ref.] | ? | ? | ? | ? |
| DAA era (≥2014) | **0.32 (0.31-0.32)** | **0.37 (0.37-0.38)** | **0.36 (0.35-0.37)** | **0.39 (0.38-0.41)** |

### Time-to confirmatory RNA testing

Predictors displaying the largest effect on the rate of confirmatory RNA testing included birth year, liver disease severity, coinfection with HIV or HBV, substance use disorder, immigration status, and treatment era (**Figure 3a**). Based on the hazard model (**Table 4**), RNA testing rate was higher among younger individuals born after 1965 (aHR: 1.26, 95%CI, 1.23-1.29 vs. baby-boomers), HIV-coinfected individuals (aHR: 1.34, 95%CI, 1.26-1.34), immigrants (aHR: 1.15, 95%CI, 1.12-1.18 vs. long-term residents), and those with advanced liver disease (aHR: 1.30-1.40 vs. without cirrhosis). RNA confirmation rate was also substantially greater for those initially diagnosed in the DAA era (aHR: 1.73, 95%CI, 1.70-1.76 vs. pre-DAA era). In contrast, RNA testing rate was lower for oldest individuals born before 1945 (aHR: 0.66, 95%CI, 0.64-0.68 vs. baby-boomers), individuals with substance use disorder (aHR: 0.86, 95%CI, 0.85-0.88) and those facing greater social marginalisation (i.e., lower income, greater housing instability and material deprivation).

**Table 4.** Predictors of time-to-engagement with key stages of the HCV care cascade.

| Covariates | AB Diagnosis to RNA Testing<br>N=87,958 |  | RNA Diagnosis to Treatment Initiation<br>N=69,509 |  | Treatment Initiation to SVR<br>N=33,063 |  |
| --- | --- | --- | --- | --- | --- | --- |
|  | Unadjusted<br>HR (95% CI) | Adjusted<br>HR (95% CI) | Unadjusted<br>HR (95% CI) | Adjusted<br>HR (95% CI) | Unadjusted<br>HR (95% CI) | Adjusted<br>HR (95% CI) |
| **Age (yrs.)** |  |  |  |  |  |  |
| At time of diagnosis | 1.01 (1.01-1.01) | 1.02 (1.02-1.02) | 1.00 (1.00-1.00) | 1.02 (1.02-1.02) | ▯ | ▯ |
| At time of treatment | ▯ | ▯ | ▯ | ▯ | 1.01 (1.00-1.01) | 1.02 (1.02-1.02) |
| **Birth year** |  |  |  |  |  |  |
| 1945-1965 [ref.] | ▯ | ▯ | ▯ | ▯ | ▯ | ▯ |
| <1945 | 1.08 (1.05-1.11) | 0.66 (0.64-0.68) | 0.63 (0.60-0.66) | 0.38 (0.36-0.40) | 0.77 (0.73-0.82) | 0.57 (0.53-0.61) |
| >1965 | 0.95 (0.93-0.96) | 1.26 (1.23-1.29) | 1.01 (0.99-1.04) | 1.34 (1.29-1.39) | 0.94 (0.91-0.96) | 1.24 (1.19-1.30) |
| **Sex** |  |  |  |  |  |  |
| Male [ref.] | ▯ | ▯ | ▯ | ▯ | ▯ | ▯ |
| Female | 1.05 (1.04-1.07) | 1.05 (1.03-1.06) | 1.09 (1.07-1.11) | 1.10 (1.08-1.13) | 1.01 (1.01-1.01) | 1.06 (1.04-1.09) |
| **Rurality** |  |  |  |  |  |  |
| Urban [ref.] | ▯ | ▯ | ▯ | ▯ | ▯ | ▯ |
| Rural | 1.02 (1.00-1.05) | 0.96 (0.93-0.98) | 0.97 (0.93-1.00) | 0.98 (0.94-1.03) | 1.04 (0.99-1.09) | 0.96 (0.92-1.01) |
| **Neighborhood income quintile** |  |  |  |  |  |  |
| Low (q. 1-2) [ref.] | ▯ | ▯ | ▯ | ▯ | ▯ | ▯ |
| Medium (q. 3) | 1.13 (1.11-1.15) | 1.01 (0.98-1.03) | 0.96 (0.94-0.99) | 1.00 (0.96-1.03) | 1.11 (1.08-1.15) | 1.03 (0.99-1.07) |
| High (q. 4-5) | 1.07 (1.05-1.09) | 1.03 (1.01-1.06) | 0.98 (0.96-1.01) | 1.00 (0.96-1.04) | 1.06 (1.02-1.09) | 1.07 (1.03-1.12) |
| **Residential instability quintile** |  |  |  |  |  |  |
| Low (q. 1-2) [ref.] | ▯ | ▯ | ▯ | ▯ | ▯ | ▯ |
| Medium (q. 3) | 0.84 (0.83-0.86) | 0.96 (0.94-0.98) | 0.98 (0.95-1.01) | 0.96 (0.93-0.99) | 0.90 (0.87-0.92) | 0.95 (0.91-0.99) |
| High (q. 4-5) | 0.93 (0.91-0.95) | 0.91 (0.89-0.93) | 0.96 (0.93-0.99) | 0.96 (0.93-0.98) | 0.94 (0.91-0.98) | 0.93 (0.90-0.96) |
| **Material deprivation quintile** |  |  |  |  |  |  |
| Low (q. 1-2) [ref.] | ▯ | ▯ | ▯ | ▯ | ▯ | ▯ |
| Medium (q. 3) | 0.87 (0.85-0.88) | 0.97 (0.95-0.99) | 1.05 (1.03-1.08) | 1.06 (1.02-1.10) | 0.93 (0.90-0.96) | 1.04 (0.99-1.09) |
| High (q. 4-5) | 0.94 (0.92-0.96) | 0.95 (0.93-0.97) | 1.05 (1.02-1.09) | 1.07 (1.03-1.10) | 1.00 (0.96-1.04) | 1.02 (0.98-1.06) |
| **Ethnic concentration quintile** |  |  |  |  |  |  |
| Low (q. 1-2) [ref.] | ▯ | ▯ | ▯ | ▯ | ▯ | ▯ |
| Medium (q. 3) | 0.98 (0.96-0.99) | 0.97 (0.95-0.99) | 1.04 (1.01-1.06) | 0.98 (0.95-1.01) | 0.96 (0.93-0.99) | 1.00 (0.96-1.04) |
| High (q. 4-5) | 0.97 (0.95-0.99) | 0.96 (0.94-0.98) | 0.98 (0.95-1.01) | 1.01 (0.98-1.03) | 0.98 (0.94-1.01) | 1.02 (0.98-1.06) |
| **Immigrant status** |  |  |  |  |  |  |
| Long-term resident [ref.] | ▯ | ▯ | ▯ | ▯ | ▯ | ▯ |
| Immigrant | 1.21 (1.19-1.24) | 1.15 (1.12-1.18) | 1.36 (1.32-1.40) | 1.37 (1.33-1.42) | 1.01 (0.97-1.04) | 1.09 (1.05-1.13) |
| **Substance use disorder\*** |  |  |  |  |  |  |
| No [ref.] | ▯ | ▯ | ▯ | ▯ | ▯ | ▯ |
| Yes | 0.92 (0.91-0.94) | 0.86 (0.85-0.88) | 0.57 (0.55-0.59) | 1.02 (0.99-1.04) | 0.95 (0.93-0.98) | 0.90 (0.87-0.92) |
| **HIV positivity\*** |  |  |  |  |  |  |
| No [ref.] | ▯ | ▯ | ▯ | ▯ | ▯ | ▯ |
| Yes | 1.20 (1.12-1.28) | 1.34 (1.25-1.44) | 1.04 (0.95-1.14) | 1.10 (1.00-1.21) | 0.87 (0.79-0.96) | 0.96 (0.87-1.05) |
| **HBsAg positivity\*** |  |  |  |  |  |  |
| No [ref.] | ▯ | ▯ | ▯ | ▯ | ▯ | ▯ |
| Yes | 1.25 (1.15-1.36) | 1.17 (1.07-1.28) | 2.48 (2.08-2.95) | 1.05 (0.90-1.22) | 0.75 (0.66-0.85) | 0.80 (0.70-0.91) |
| **Total Number of ADGs (N.)\*** | 1.01 (1.01-1.01) | 1.01 (1.01-1.01) | 1.00 (1.00-1.00) | 1.00 (1.00-1.00) | 0.98 (0.97-0.98) | 0.98 (0.98-0.99) |
| **Liver disease\*** |  |  |  |  |  |  |
| No cirrhosis [ref.] | ▯ | ▯ | ▯ | ▯ | ▯ | ▯ |
| Compensated cirrhosis | 1.36 (1.32-1.40) | 1.30 (1.26-1.34) | 1.07 (1.03-1.11) | 1.07 (1.03-1.11) | 1.22 (1.18-1.26) | 1.15 (1.11-1.18) |
| Decompensated cirrhosis | 1.35 (1.32-1.39) | 1.37 (1.33-1.40) | 1.09 (1.05-1.13) | 1.15 (1.11-1.19) | 0.94 (0.90-0.98) | 1.06 (1.01-1.11) |
| Hepatocellular carcinoma | 1.45 (1.40-1.51) | 1.40 (1.35-1.46) | 0.98 (0.93-1.03) | 1.02 (0.97-1.07) | 0.92 (0.86-0.99) | 1.01 (0.94-1.08) |
| **Retreatment with HCV antivirals (ever)** |  |  |  |  |  |  |
| No [ref.] | ▯ | ▯ | ▯ | ▯ | ▯ | ▯ |
| Yes | ▯ | ▯ | ▯ | ▯ | 0.48 (0.46-0.50) | 0.56 (0.53-0.59) |
| **HCV diagnosis/treatment (first time)** |  |  |  |  |  |  |
| Pre-DAA era (<2014) [ref.] | ▯ | ▯ | ▯ | ▯ | ▯ | ▯ |
| DAA era (≥2014) | 1.79 (1.76-1.82) | 1.73 (1.70-1.76) | 2.14 (2.07-2.21) | 1.95 (1.88-2.01) | 2.48 (2.41-2.55) | 2.16 (2.09-2.23) |

\*Baseline covariates are collected at the time of origin for each model (i.e., at the time of HCV diagnosis for RNA testing and treatment initiation and the time of treatment initiation for SVR). A look-back period of one year was used to collect ADGs. Bold values indicate statistical significance at $p < 0.05$ . *Abbreviations:* SVR: sustained virologic response; AB: antibody; RNA: ribonucleic acid; HCV: hepatitis c virus; DAA: direct-acting antiviral; ADG: aggregated diagnostic groups; HBsAg: hepatitis b surface antigen; HIV: Human immunodeficiency virus; HR: hazard ratio; CI: confidence interval; q: quintile; N: number of observations.

### Time-to treatment initiation

The predictors most strongly associated with the timing of HCV treatment initiation following confirmation of RNA positivity included birth year, sex, treatment era, and liver disease severity (**Figure 3b**). The rate of treatment initiation was higher for younger individuals (aHR: 1.34, 95%CI, 1.29-1.39 vs. baby-boomers), females (aHR: 1.10, 95%CI, 1.08-1.13 vs. males), immigrants (aHR: 1.37, 95%CI, 1.33-1.42 vs. long-term residents), as well as those diagnosed with cirrhosis (aHR: 1.07, 95%CI, 1.03-1.11 and aHR: 1.15, 95%CI, 1.11-1.19, for compensated and DC respectively vs. without cirrhosis) (**Table 4**). Treatment initiation was also higher for those diagnosed in the DAA era (aHR: 1.95, 95%CI, 1.88-2.01 vs. pre-DAA era). In contrast, older individuals (aHR: 0.38, 95%CI, 0.36-0.40 vs. baby-boomers) experienced lower rates of treatment as did individuals facing higher levels of housing instability (aHR: 0.96, 95%CI, 0.93-0.99 vs. low instability). However, material deprivation was associated with higher rates of treatment (aHR: 1.07, 95%CI, 1.03-1.10 vs. low deprivation), and indeed most treated individuals in this study acquired HCV antivirals through public drug programs for those in financial need.

### Time-to SVR

Predictors most strongly associated with SVR included birth year, treatment era, HBV-coinfection, and retreatment (**Figure 3c**). SVR rate was significantly higher among younger individuals (aHR: 1.24, 95%CI, 1.19-1.30 vs. baby-boomers), females (aHR: 1.06, 95%CI, 1.04-1.08 vs. males), those residing in higher income settings (aHR: 1.07, 95%CI, 1.03-1.12 vs. lowest), immigrants (aHR: 1.09, 95%CI, 1.05-1.13 vs. long-term residents), those initiating treatment in the DAA-era (aHR: 2.16, 95%CI, 2.09-2.23 vs. pre-DAA era), and those with compensated cirrhosis (aHR: 1.15 95%CI, 1.11-1.18 vs. without cirrhosis) (**Table 4**). In contrast, rate of SVR achievement was lower for older subjects born before 1945 (aHR: 0.57, 95%CI, 0.53-0.61 vs. baby-boomers), those with HBV-coinfection (aHR: 0.80, 95%CI, 0.70-0.91) substance use (aHR: 0.90, 95%CI, 0.87-0.92), residential instability (aHR: 0.93, 95%CI, 0.90-0.96 vs. lowest instability) and those requiring retreatment with HCV antivirals (aHR: 0.56, 95%CI, 0.53-0.59).

## Discussion

We conducted a population-based cohort stud to describe the current state of HCV care cascade and to explore predictors of engagement with HCV care in the province. We found that 88% of Ontario residents with an antibody diagnosis of HCV received confirmatory RNA testing and of these, 62% of tested positive for RNA. However, only 53% of individuals testing RNA positive initiated treatment, and 76% of those treated achieved SVR, while ∼1% experienced reinfection or relapse by the end of 2018. We also found that treatment initiation appeared to be the main gap in the care cascade across different risk populations.

Our findings were largely congruent with previous work by Yasseen et. al. that evaluated the HCV care cascade in Ontario primarily in the pre-DAA era (1999-2014) but showed improvement in treatment initiation and SVR achievement in the DAA-era. Our findings were also consistent with the earlier study in showing higher engagement with the care cascade among immigrants relative to long-term residents regardless of HCV endemicity status of immigrants’ country of origin (2). Findings of the current study were also consistent with clinic-based Canadian data showing treatment initiation and SVR levels to be comparable between immigrants and non-immigrants (15).

Moreover, Ontario’s population-level care cascade is also consistent with a recent (2018) study from British Colombia (BC), where treatment initiation was also identified as a major gap (16). Although, compared to BC, the level of confirmatory RNA testing in Ontario was slightly higher (88% vs. 83%) while RNA positivity was lower (62% vs. 72%). Moreover, treatment initiation also appeared to be lower in Ontario (53% vs. 61%). This is likely related to inter-provincial differences with respect to DAA reimbursement restrictions between 2014 and 2018. Furthermore, treatment levels in Ontario are also likely to be underestimated due to missing data on privately funded HCV antiviral dispensations. Finally, while based on the current study SVR levels in Ontario showed a marked improvement relative to earlier data from the pre-DAA era (76% vs. 45%), the 2018 SVR levels in Ontario tended to be lower relative to BC (76% vs. 90%). Such differences in SVR could result from methodological differences in terms of definition of SVR and the application of backfilling and/or inflation factors to account for missing data on treatment initiation. Moreover, given the cross-sectional nature of care cascade studies, SVR measurement is likely subject to right censoring due to incomplete treatment duration and thus is likely to underestimate SVR.

With respect to our secondary objectives, we found that following adjustments for multiple covariates, individuals facing unstable housing or substance use to display significantly lower rates of engagement with HCV RNA testing despite experiencing significantly higher rates of HCV antibody and RNA test positivity. Given these findings the use of reflex testing to reduce loss between antibody and RNA testing and implementation of HCV screening through emergency departments that may prove useful in reaching high-risk populations may be warranted. Indeed, screening programmes implemented through emergency departments have been shown to be cost-effective options along with the use of point-of-care assays that can facilitate HCV RNA diagnosis (17–19).

Our study has limitations. First, our analysis likely underestimates treatment levels since HCV dispensation records are based on Ontario’s public drug plans. However, to account for individuals who may have received treatment outside of the provincial drug plan (i.e., through private drug plans, out-of-pocket or out-of-province), the treatment initiation and SVR stages were supplemented by those who had a final negative test result indicative of having achieved SVR, excluding those who met the definition of acute resolved infections. Second, because PHO laboratory data only cover HCV testing from 1999 to 2018, we lack data on testing records prior to 1999 and those initially diagnosed out-of-province, although backfilling will likely capture some of those cases. As well, individuals who moved and continued treatment outside of Ontario will not be captured in the current estimates. Third, given that the IRCC dataset starts in 1985, individuals immigrating to Canada before 1985 are classified as long-term residents in line with previous studies (2). Finally, given the cross-sectional nature of the care cascade study, SVR proportion is subject to right censoring. Similarly, with respect to reinfection/relapse, the current study presents a snapshot of the number of individuals who may need to reengage with HCV care as of December 2018; however, since individuals are not routinely tested for reinfection, reinfection/relapse level is likely to be an underestimate.

Our study has several strengths. We assembled a large population-based sample of individuals undergoing testing for HCV in the province of Ontario, Canada’s most populous jurisdiction, and used linked databases to describe the DAA-era state of HCV care in the province. We used cause-specific hazard models to identify clinical and socio-economic factors associated with the rate of engagement with each step of the cascade and explored predictors of HCV test positivity which will be useful in informing scaling-up of HCV screening and linkage-to-care strategies and identifying key populations. Finally, the availability of a real-world population-based sample allows for generalizable findings for provincial policymaking and should act as a benchmark to help assess the impact of elimination efforts and to monitor progress.

In conclusion, treatment initiation appears as a major gap in care for most groups. Given that short delays in DAA treatment can have profound impacts on the morbidity and mortality of individuals living with hepatitis C, rapid linkage to curative treatment specifically for priority populations will be needed (20). More recently, HCV testing rates have stagnated due to the COVID-19 pandemic (21). Therefore, innovative expansion of HCV screening and linkage-to-care initiatives is also warranted. Particularly, in Ontario, our findings suggest that improvements in access to HCV care for those with a history of substance use and social marginalisation will be needed to equitably close the gaps in the HCV care cascade. Future work will be needed to assess feasibility and inform implementation of such programs.

## Data Availability

The dataset from this study is held securely in coded form at ICES. While legal data sharing agreements between ICES and data providers (e.g., healthcare organizations and government) prohibit ICES from making the dataset publicly available, access may be granted to those who meet pre-specified criteria for confidential access, available at www.ices.on.ca/DAS. The full dataset creation plan and underlying analytic code are available from the authors upon request, understanding that the computer programs may rely upon coding templates or macros that are unique to ICES and are therefore either inaccessible or may require modification.

## Acknowledgements

Parts or whole of this material are based on data and/or information compiled and provided by Immigration, Refugees and Citizenship Canada (IRCC) current to May 2017. However, the analyses, conclusions, opinions, and statements expressed in the material are those of the author(s), and not necessarily those of IRCC. We thank IQVIA Solutions Canada Inc. for use of their Drug Information File. Parts of this material are based on data and/or information compiled and provided by CIHI. However, the analyses, conclusions, opinions, and statements expressed in the material are those of the author(s), and not necessarily those of CIHI. Parts of this report are based on Ontario Registrar General (ORG) information on deaths, the original source of which is Service Ontario. The views expressed therein are those of the author and do not necessarily reflect those of ORG or the Ministry of Government Services. We thank Dr. Andrew Mendlowitz (Toronto Center for Liver Disease, University Health Network, Toronto, ON) for methodological guidance and feedback.

## Supplemental Tables and Figures

**Supplemental Table 1.**
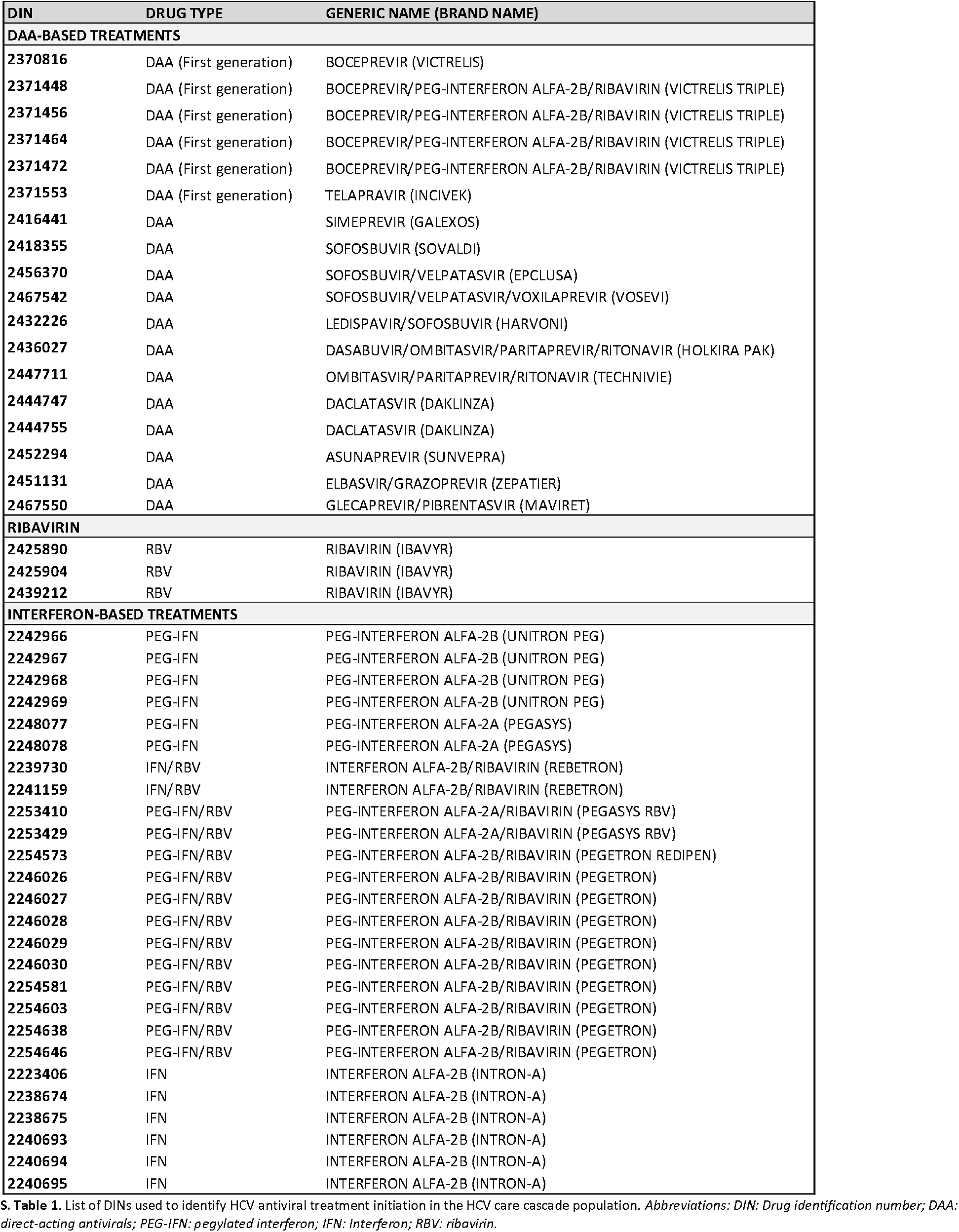
Drug identification numbers used to identify HCV antiviral treatment.

**Supplemental Table 2A.**
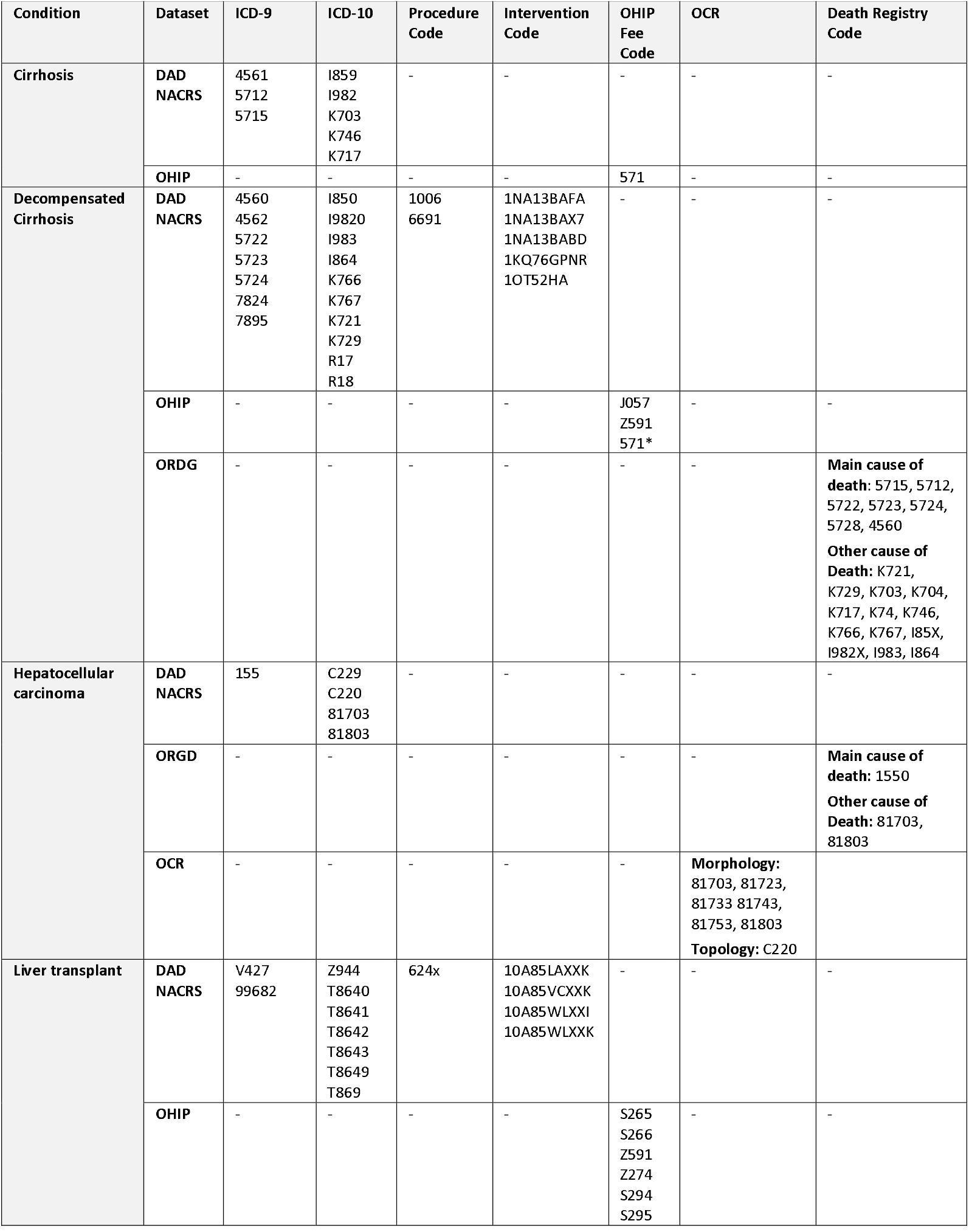
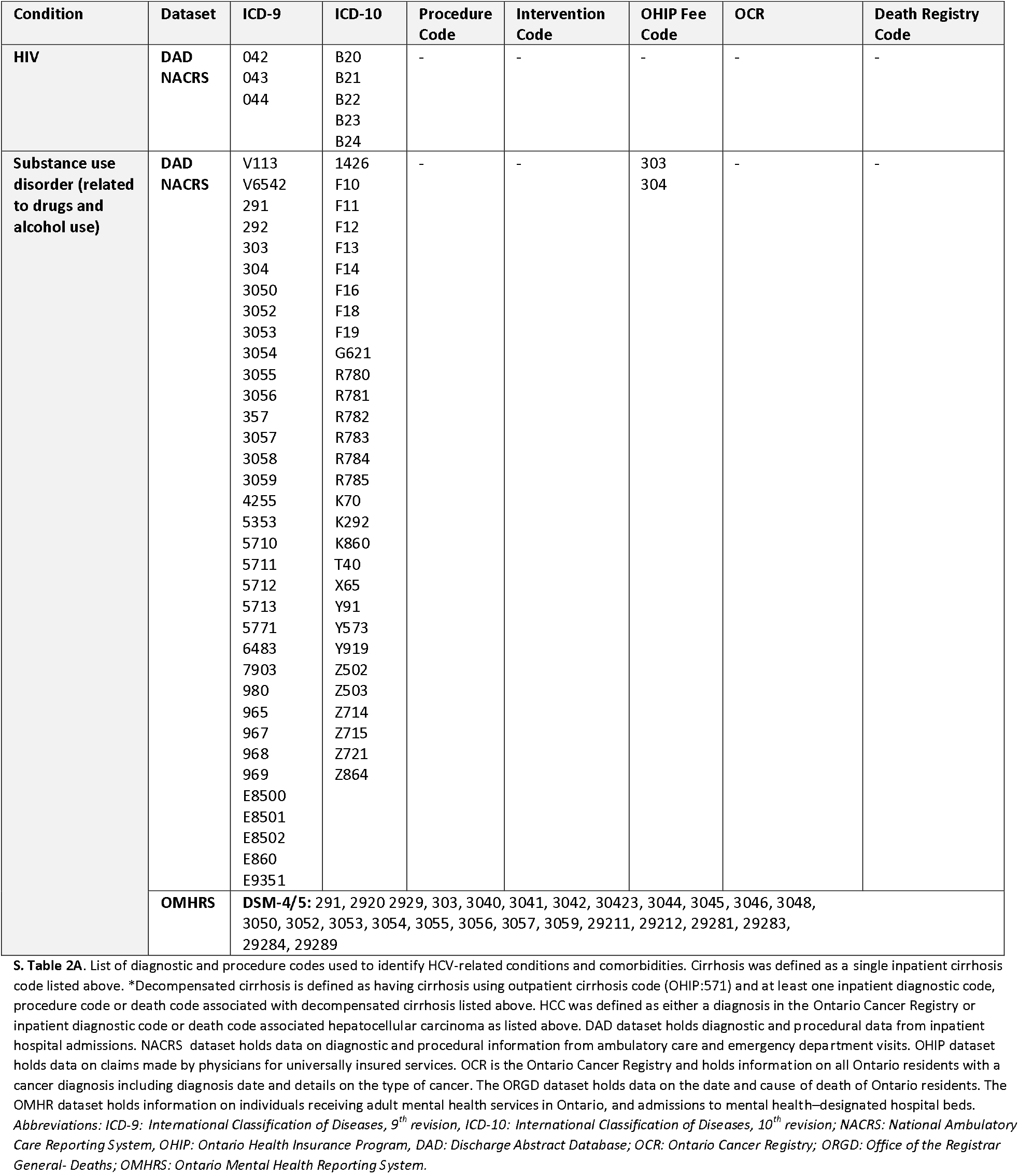
Diagnostic and procedure codes used to identify HCV-related diagnosis and comorbidities from datasets held at ICES.

**Supplemental Table 2B.**
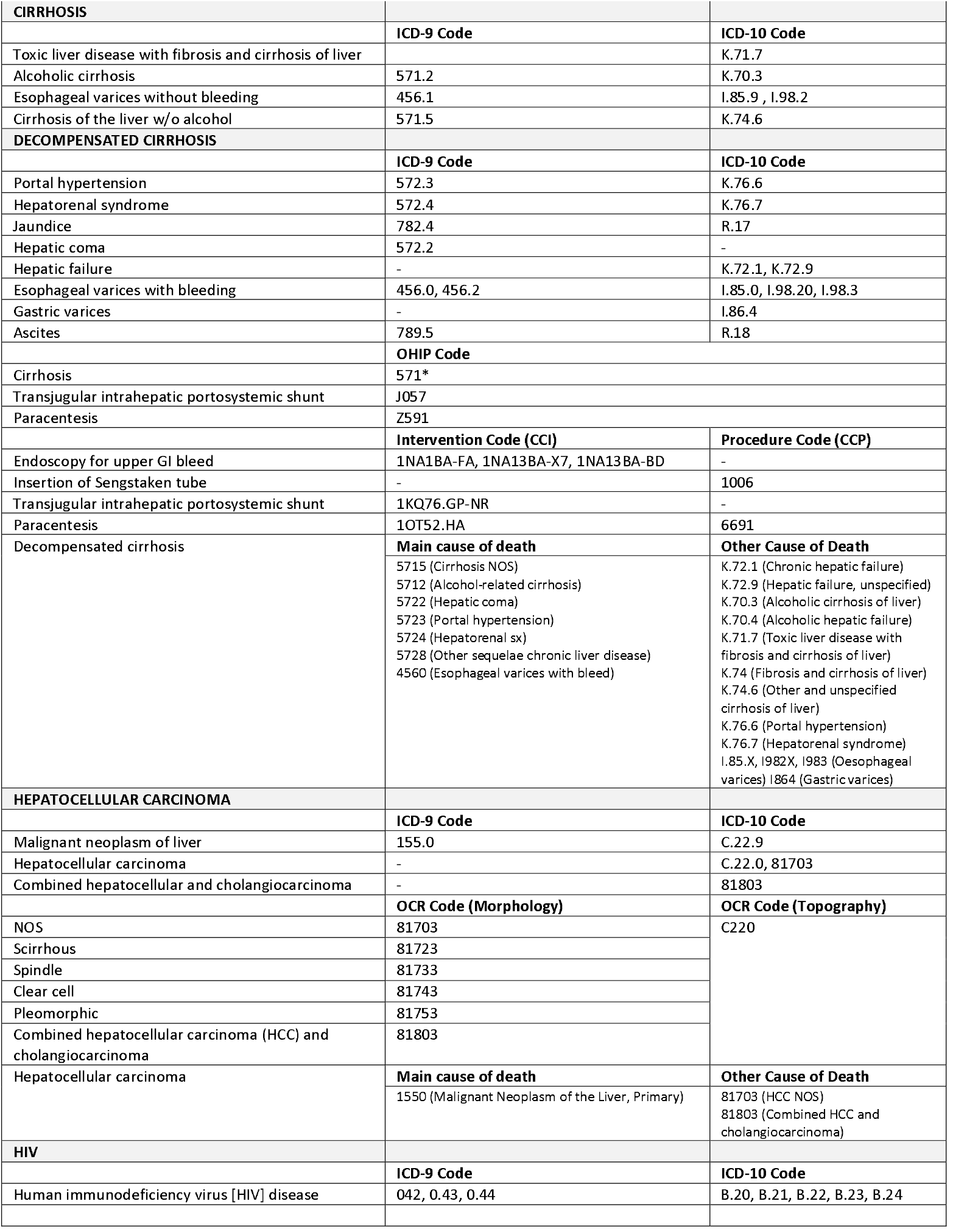
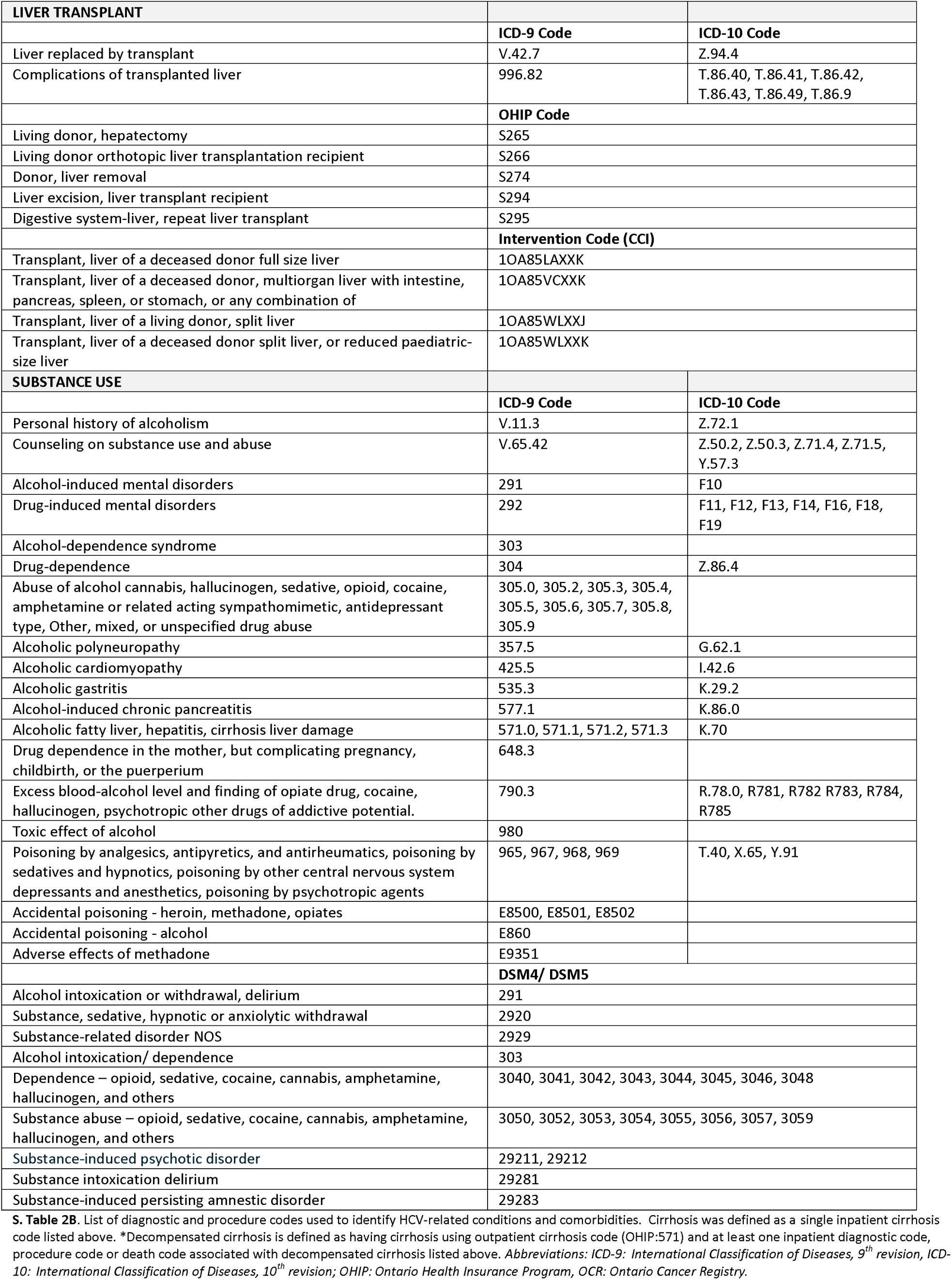
Detailed description of diagnostic, death and procedure-related codes used.

**S. Table 3.** Baseline characteristics stratified by care cascade stage.

|  | TOTAL ONTARIO POPULATION |  |  |  |  |  |  |
| --- | --- | --- | --- | --- | --- | --- | --- |
|  | HCV AB<br>Positive<br>N=108,428 | RNA<br>Tested<br>N=95,002 | RNA<br>Positive<br>N=59,370 | GT<br>Tested<br>N=56,103 | Treatment<br>Initiated<br>N=31,616 | SVR<br>Achieved<br>N=23,950 | Reinfection or<br>Relapse<br>N=242 |
| **Age in years, mean (SD)** |  |  |  |  |  |  |  |
| Age at diagnosis | 42.6 (14.3) | 42.9 (14.2) | 43.1 (12.5) | 43.4 (12.4) | 44.3 (11.8) | 44.6 (11.7) | 42.7 (12.2) |
| Age at treatment | 51.0 (12.7) | 51.1 (12.7) | 51.1 (12.7) | 51.1 (12.7) | 51.4 (12.5) | 52.1 (11.7) | 47.8 (12.5) |
| Age at SVR | 54.6 (12.2) | 54.6 (12.2) | 54.6 (12.2) | 54.6 (12.2) | 54.6 (12.2) | 54.6 (12.2) | 46.2 (12.1) |
| **Birth cohort, N (%)** |  |  |  |  |  |  |  |
| <1945 | 5,537 (5.2) | 4,733 (5) | 2,789 (4.6) | 2,636 (4.7) | 1,431 (4.5) | 1,074 (4.5) | 13 (5.4) |
| 1945-1965 | 52,750 (48.6) | 47,152 (49.6) | 32,447 (54.7) | 31,082 (55.4) | 19,307 (61.1) | 15,172 (63.3) | 129 (53.3) |
| >1965 | 50,141 (46.2) | 43,117 (45.4) | 24,134 (40.7) | 22,385 (39.9) | 10,878 (34.4) | 7,704 (32.2) | 100 (41.3) |
| **Male sex, N (%)** | 64,835 (60.0) | 56,240 (59.3) | 37,824 (63.8) | 35,925 (64.1) | 19,876 (62.9) | 14,965 (62.5) | 156 (64.5) |
| **Rural, N (%)** | 10,544 (9.9) | 9,231 (9.9) | 6,147 (10.5) | 5,758 (10.4) | 2,993 (9.5) | 2,240 (9.4) | 24 (10.0) |
| **Neighborhood income quintile, N (%)** |  |  |  |  |  |  |  |
| 1st quintile (lowest) | 35,297 (33.5) | 30,416 (32.6) | 20,219 (34.8) | 19,108 (34.8) | 10,562 (33.7) | 7,701 (32.5) | 90 (37.8) |
| 2nd quintile | 23,346 (22.1) | 20,617 (22.1) | 13,199 (22.7) | 12,493 (22.7) | 7,204 (23.0) | 5,469 (23.0) | 63 (26.5) |
| 3rd quintile | 18,674 (17.7) | 16,592 (17.8) | 10,122 (17.4) | 9,608 (17.5) | 5,509 (17.6) | 4,238 (17.9) | 32 (13.4) |
| 4th quintile | 15,554 (14.7) | 14,041 (15.1) | 8,254 (14.2) | 7,816 (14.2) | 4,537 (14.5) | 3,592 (15.1) | 30 (12.6) |
| 5th quintile (highest) | 12,715 (12.0) | 11,569 (12.4) | 6,315 (10.9) | 5,935 (10.8) | 3,506 (11.2) | 2,728 (11.5) | 23 (9.7) |
| **Residential instability quintile, N (%)** |  |  |  |  |  |  |  |
| 1st quintile (lowest) | 13,727 (13.4) | 12,655 (13.9) | 6,437 (11.3) | 6,052 (11.3) | 3,705 (12.0) | 3,017 (12.9) | 27 (11.4) |
| 2nd quintile | 13,865 (13.4) | 12,533 (13.7) | 7,368 (13.0) | 6,967 (13.0) | 4,103 (13.3) | 3,212 (13.8) | 26 (11.0) |
| 3rd quintile | 16,148 (15.6) | 14,429 (15.8) | 9,021 (15.9) | 8,560 (15.9) | 4,879 (15.9) | 3,753 (16.1) | 33 (14.0) |
| 4th quintile | 22,950 (22.2) | 19,948 (21.9) | 13,143 (23.2) | 12,413 (23.1) | 7,072 (23.0) | 5,289 (22.7) | 66 (28.0) |
| 5th quintile (highest) | 36,510 (35.4) | 31,657 (34.7) | 20,767 (36.6) | 19,702 (36.7) | 10,996 (35.8) | 8,027 (34.5) | 84 (35.6) |
| **Material deprivation quintile, N (%)** |  |  |  |  |  |  |  |
| 1st quintile (lowest) | 13,120 (12.7) | 11,945 (13.1) | 6,527 (11.5) | 6,143 (11.5) | 3,573 (11.6) | 2,737 (11.7) | 17 (7.2) |
| 2nd quintile | 14,877 (14.4) | 13,412 (14.7) | 7,897 (13.9) | 7,468 (13.9) | 4,341 (14.1) | 3,425 (14.7) | 30 (12.7) |
| 3rd quintile | 17,778 (17.2) | 15,933 (17.5) | 9,756 (17.2) | 9,246 (17.2) | 5,427 (17.6) | 4,205 (18.0) | 38 (16.1) |
| 4th quintile | 21,794 (21.1) | 19,256 (21.1) | 12,155 (21.4) | 11,512 (21.4) | 6,622 (21.5) | 5,046 (21.7) | 57 (24.2) |
| 5th quintile (highest) | 35,631 (34.5) | 30,676 (33.6) | 20,401 (36.0) | 19,325 (36.0) | 10,792 (35.1) | 7,885 (33.8) | 94 (39.8) |
| **Ethnic concentration quintile, N (%)** |  |  |  |  |  |  |  |
| 1st quintile (lowest) | 17,031 (16.5) | 14,838 (16.3) | 10,152 (17.9) | 9,599 (17.9) | 5,194 (16.9) | 3,867 (16.6) | 50 (21.7) |
| 2nd quintile | 17,648 (17.1) | 15,933 (16.9) | 10,257 (18.1) | 9,686 (18.0) | 5,371 (17.5) | 4,016 (17.2) | 50 (21.2) |
| 3rd quintile | 18,213 (17.6) | 19,050 (17.6) | 10,355 (18.3) | 9,771 (18.2) | 5,456 (17.7) | 4,100 (17.6) | 35 (14.8) |
| 4th quintile | 21,762 (21.1) | 19,208 (21.1) | 11,820 (20.8) | 11,238 (20.9) | 6,390 (20.8) | 4,840 (20.8) | 46 (19.5) |
| 5th quintile (highest) | 28,546 (27.7) | 25,742 (28.2) | 14,152 (24.9) | 13,400 (25.0) | 8,344 (27.1) | 6,475 (27.8) | 55 (23.3) |
| **Immigrant N (%)** | 16,851 (15.5) | 15,596 (16.4) | 7,836 (13.2) | 7,424 (13.2) | 4,872 (15.4) | 3,892 (16.3) | 33 (13.6) |
| **Substance use disorder\*, N (%)** | 31,669 (29.2) | 26,418 (27.8) | 19,463 (32.8) | 18,376 (32.8) | 11,862 (37.5) | 8,257 (34.5) | 125 (51.7) |
| **HIV positivity\*, N (%)** | 1,170 (1.1) | 1,101 (1.2) | 595 (1.0) | 595 (1.0) | 620 (2.0) | 435 (1.8) | 7 (2.9) |
| **HBV status\*, N (%)** |  |  |  |  |  |  |  |
| HBV antibody positive (ever) | 12,406 (11.4) | 11,508 (12.1) | 7,012 (11.8) | 6,456 (11.5) | 3,614 (11.4) | 2,666 (11.1) | 34 (14.0) |
| HBV antigen positive | 1,615 (1.5) | 1,543 (1.6) | 373 (0.6) | 337 (0.6) | 355 (1.1) | 229 (1.0) | 1-5 (≤2.1) |
| **Aggregated diagnosis group categories\*, N (%)** |  |  |  |  |  |  |  |
| 0-3 ADGs | 41,265 (38.2) | 35,640 (37.6) | 22,715 (38.3) | 21,466 (38.3) | 8,792 (27.9) | 6,926 (28.9) | 55 (22.7) |
| 4-7 ADGs | 43,278 (40.0) | 38,444 (40.5) | 23,745 (40.1) | 22,478 (40.1) | 14,204 (44.9) | 10,781 (45) | 101 (41.7) |
| 8-10 ADGs | 15,544 (14.4) | 13,786 (14.5) | 8,541 (14.4) | 8,065 (14.4) | 5,636 (17.8) | 4,119 (17.2) | 58 (24.0) |
| >11 ADGs | 7,995 (7.4) | 6,971 (7.4) | 4,281 (7.2) | 4,022 (7.2) | 2,984 (9.4) | 2,124 (8.9) | 28 (11.6) |
| **Liver disease categories\*, N (%)** |  |  |  |  |  |  |  |
| Non-cirrhotic | 95,911 (88.5) | 83,248 (87.6) | 50,205 (84.6) | 47,278 (84.3) | 21,343 (67.4) | 15,794 (66.0) | 172 (71) |
| Compensated cirrhosis | 5,754 (5.3) | 5,325 (5.6) | 4,096 (6.9) | 3,923 (7.0) | 7,515 (23.8) | 6,067 (25.3) | 36 (14.9) |
| Decompensated cirrhosis | 4,776 (4.4) | 4,540 (4.8) | 3,505 (5.9) | 3,391 (6.0) | 1,822 (5.8) | 1,350 (5.6) | 21 (8.7) |
| Hepatocellular carcinoma | 1,987 (1.8) | 1,889 (2) | 1,564 (2.6) | 1,511 (2.7) | 936 (3.0) | 739 (3.1) | 13 (5.4) |
| **HCV genotype, N (%)** |  |  |  |  |  |  |  |
| Genotype 1 | 34,471 (63.0) | 34,471 (63.0) | 34,471 (63.0) | 34,471 (63.0) | 18,453 (62.7) | 14,251 (63.4) | 122 (55) |
| Genotype 2 | 5,959 (10.9) | 5,959 (10.9) | 5,959 (10.9) | 5,959 (10.9) | 3,289 (11.2) | 2,556 (11.4) | 23 (10.4) |
| Genotype 3 | 11,792 (21.6) | 11,792 (21.6) | 11,792 (21.6) | 11,792 (21.6) | 6,360 (21.6) | 4,675 (20.8) | 72 (32.4) |
| Genotype 4 | 1,149 (2.1) | 1,149 (2.1) | 1,149 (2.1) | 1,149 (2.1) | 652 (2.2) | 484 (2.2) | 1-5 (≤2.1) |
| Genotype 5 | 87 (0.2) | 87 (0.2) | 87 (0.2) | 87 (0.2) | 44 (0.1) | 34 (0.2) | 0 (.) |
| Genotype 6 | 375 (0.7) | 375 (0.7) | 375 (0.7) | 375 (0.7) | 242 (0.8) | 202 (0.9) | 1-5 (≤2.3) |
| Coinfected | 861 (1.6) | 861 (1.6) | 861 (1.6) | 861 (1.6) | 403 (1.4) | 293 (1.3) | 1-5 (≤2.3) |
| **HCV diagnosis (first time), N (%)** |  |  |  |  |  |  |  |
| Pre-DAA era (<2014) | 77,859 (71.8) | 68,326 (71.9) | 46,956 (79.1) | 44,598 (79.5) | 26,425 (83.6) | 20,601 (86.0) | 210 (86.8) |
| DAA era (≥2014) | 30,569 (28.2) | 26,676 (28.1) | 12,414 (20.9) | 11,505 (20.5) | 5,191 (16.4) | 3,349 (14.0) | 32 (13.2) |
| **Treatment initiation (first time), N (%)** |  |  |  |  |  |  |  |
| Pre-DAA era (<2014) | 12,716 (40.2) | 12,716 (40.2) | 12,716 (40.2) | 12,320 (39.9) | 12,716 (40.2) | 9,982 (41.7) | 152 (62.8) |
| DAA era (≥2014) | 18,900 (59.8) | 18,900 (59.8) | 18,900 (59.8) | 18,532 (60.1) | 18,900 (59.8) | 13,968 (58.3) | 90 (37.2) |
| **Drug plan, N (%)** |  |  |  |  |  |  |  |
| Low income (Trillium or Special Drugs Program) | 7,009 (22.2) | 7,009 (22.2) | 7,009 (22.2) | 7,009 (22.2) | 7,009 (22.2) | 4,993 (20.9) | 54 (22.3) |
| Low income (Ontario Works) | 4,091 (12.9) | 4,091 (12.9) | 4,091 (12.9) | 4,091 (12.9) | 4,091 (12.9) | 2,397 (10.0) | 68 (28.0) |
| Senior (Senior and Low-Income Senior) | 2,703 (8.6) | 2,703 (8.6) | 2,703 (8.6) | 2,703 (8.6) | 2,703 (8.6) | 1,945 (8.1) | 19 (7.9) |
| Long-term care or home care | 176 (0.6) | 176 (0.6) | 176 (0.6) | 176 (0.6) | 176 (0.6) | 61 (0.3) | 0 (.) |
| OHIP+ (for 24 years and younger) | 145 (0.5) | 145 (0.5) | 145 (0.5) | 145 (0.5) | 145 (0.5) | 43 (0.2) | 0 (.) |
| Other public plan | 8,003 (25.3) | 8,003 (25.3) | 8,003 (25.3) | 8,003 (25.3) | 8,003 (25.3) | 5,031 (21.0) | 101 (41.7) |
| Unknown (Presumed private plan) | 9476 (30.0) | 9476 (30.0) | 9476 (30.0) | 9476 (30.0) | 9476 (30.0) | 9476 (39.6) | 0 (.) |

**S.Table 4a.** Baseline characteristics stratified by birth year (birth year<1945)

|  | BIRTH YEAR <1945 |  |  |  |  |  |  |
| --- | --- | --- | --- | --- | --- | --- | --- |
|  | HCV AB<br>Positive<br>N=5,537 | RNA<br>Tested<br>N=4,733 | RNA<br>Positive<br>N=2,789 | GT<br>Tested<br>N=2,636 | Treatment<br>Initiated<br>N=1,431 | SVR<br>Achieved<br>N=1,074 | Reinfection or<br>Relapse<br>N=13 |
| **Age in years, mean (SD)** |  |  |  |  |  |  |  |
| Age at diagnosis | 70.5 (8.0) | 70.2 (7.9) | 68.2 (7.5) | 68.1 (7.5) | 66.6 (7.0) | 66.4 (6.9) | 67.5 (6.6) |
| Age at treatment | 74.4 (6.7) | 74.3 (6.7) | 74.3 (6.7) | 74.4 (6.7) | 74.0 (6.7) | 74.7 (6.1) | 71.3 (7.0) |
| Age at SVR | 76.9 (4.7) | 76.9 (4.7) | 76.9 (4.7) | 76.9 (4.7) | 76.9 (4.7) | 76.9 (4.7) | 81.0 (2.8) |
| **Male sex, N (%)** | 2,535 (45.9) | 2,185 (46.3) | 1,259 (45.2) | 1,192 (45.3) | 663 (46.3) | 489 (45.5) | 7 (53.8) |
| **Rural, N (%)** | 263 (4.8) | 235 (5) | 122 (4.4) | 112 (4.3) | 61 (4.3) | 48 (4.5) | 1-5 (≤38.5) |
| **Neighborhood income quintile, N (%)** |  |  |  |  |  |  |  |
| 1st quintile (lowest) | 1,423 (25.9) | 1,185 (25.2) | 713 (25.8) | 670 (25.7) | 339 (23.8) | 250 (23.3) | 1-5 (≤38.5) |
| 2nd quintile | 1,181 (21.5) | 1,008 (21.5) | 619 (22.4) | 578 (22.1) | 310 (21.7) | 235 (22.0) | 1-5 (≤38.5) |
| 3rd quintile | 1,018 (18.6) | 861 (18.3) | 521 (18.8) | 503 (19.2) | 269 (18.9) | 199 (18.6) | 1-5 (≤38.5) |
| 4th quintile | 962 (17.6) | 836 (17.8) | 481 (17.4) | 460 (17.6) | 271 (19.0) | 209 (19.6) | 1-5 (≤38.5) |
| 5th quintile (highest) | 897 (16.4) | 806 (17.2) | 431 (15.6) | 404 (15.4) | 237 (16.6) | 176 (16.5) | 1-5 (≤38.5) |
| **Residential instability quintile, N (%)** |  |  |  |  |  |  |  |
| 1st quintile (lowest) | 1,161 (21.3) | 1,010 (21.7) | 587 (21.5) | 557 (21.6) | 309 (22.0) | 229 (21.7) | 6 (46.2) |
| 2nd quintile | 893 (16.5) | 766 (16.5) | 438 (16.0) | 410 (15.8) | 244 (17.3) | 186 (17.6) | 1-5 (≤38.5) |
| 3rd quintile | 867 (16.0) | 762 (16.4) | 458 (16.7) | 441 (17.0) | 230 (16.3) | 171 (16.1) | 1-5 (≤38.5) |
| 4th quintile | 1,015 (18.7) | 853 (18.3) | 520 (19.0) | 484 (18.7) | 281 (19.9) | 212 (20.0) | 1-5 (≤38.5) |
| 5th quintile (highest) | 1,491 (27.5) | 1,262 (27.1) | 732 (26.8) | 695 (26.9) | 345 (24.5) | 261 (24.6) | 1-5 (≤38.5) |
| **Material deprivation quintile, N (%)** |  |  |  |  |  |  |  |
| 1st quintile (lowest) | 933 (17.2) | 817 (17.5) | 459 (16.8) | 436 (16.8) | 241 (17.2) | 173 (16.4) | 1-5 (≤38.5) |
| 2nd quintile | 909 (16.7) | 801 (17.2) | 449 (16.4) | 424 (16.4) | 250 (17.7) | 193 (18.2) | 1-5 (≤38.5) |
| 3rd quintile | 1,010 (18.6) | 859 (18.5) | 507 (18.5) | 485 (18.7) | 274 (19.4) | 208 (19.6) | 0 (.) |
| 4th quintile | 1,112 (20.5) | 968 (20.8) | 568 (20.8) | 537 (20.8) | 282 (20) | 213 (20.1) | 1-5 (≤38.5) |
| 5th quintile (highest) | 1,463 (27) | 1,208 (26) | 752 (27.5) | 705 (27.3) | 362 (25.7) | 272 (25.7) | 1-5 (≤38.5) |
| **Ethnic concentration quintile, N (%)** |  |  |  |  |  |  |  |
| 1st quintile (lowest) | 535 (9.8) | 476 (10.3) | 238 (8.6) | 221 (8.6) | 122 (8.7) | 85 (8) | 0 (.) |
| 2nd quintile | 580 (10.7) | 509 (10.9) | 284 (10.4) | 270 (10.4) | 154 (10.9) | 123 (11.6) | 1-5 (≤38.5) |
| 3rd quintile | 789 (14.5) | 700 (15) | 385 (14.1) | 364 (14.1) | 207 (14.7) | 156 (14.7) | 1-5 (≤38.5) |
| 4th quintile | 1,289 (23.8) | 1,099 (23.6) | 680 (24.9) | 650 (25.1) | 357 (25.3) | 278 (26.3) | 1-5 (≤38.5) |
| 5th quintile (highest) | 2,234 (41.2) | 1,869 (40.2) | 1,148 (42) | 1,082 (41.8) | 569 (40.4) | 417 (39.4) | 9 (69.2) |
| **Immigrant N (%)** | 1,562 (28.2) | 1,262 (26.7) | 823 (29.5) | 786 (29.8) | 383 (26.8) | 260 (24.2) | 7 (53.8) |
| **Substance use disorder\*, N (%)** | 294 (5.3) | 252 (5.3) | 124 (4.4) | 115 (4.4) | 115 (8.0) | 84 (7.8) | 0 (.) |
| **HIV positivity\*, N (%)** | 15 (0.3) | 13 (0.3) | 7 (0.3) | 7 (0.3) | 1-5 (≤0.3) | 1-5 (≤0.5) | 0 (.) |
| **HBV status\*, N (%)** |  |  |  |  |  |  |  |
| HBV antibody positive (ever) | 192 (3.5) | 179 (3.8) | 86 (3.1) | 81 (3.1) | 49 (3.4) | 1-5 (≤0.5) | 0 (.) |
| HBV antigen positive | 62 (1.1) | 59 (1.2) | 9 (0.3) | 7 (0.3) | 1-5 (≤0.3) | 1-5 (≤0.5) | 0 (.) |
| **Aggregated diagnosis group categories\*, N (%)** |  |  |  |  |  |  |  |
| 0-3 ADGs | 1,356 (24.6) | 1,106 (23.4) | 773 (27.8) | 734 (27.9) | 194 (13.5) | 144 (13.4) | 1-5 (≤38.5) |
| 4-7 ADGs | 2,351 (42.6) | 2,063 (43.7) | 1,210 (43.5) | 1,139 (43.3) | 649 (45.4) | 490 (45.6) | 7 (53.8) |
| 8-10 ADGs | 1,136 (20.6) | 973 (20.6) | 524 (18.8) | 497 (18.9) | 369 (25.8) | 279 (26) | 1-5 (≤38.5) |
| >11 ADGs | 675 (12.2) | 582 (12.3) | 276 (9.9) | 261 (9.9) | 219 (15.3) | 161 (15) | 0 (.) |
| **Liver disease categories\*, N (%)** |  |  |  |  |  |  |  |
| Non-cirrhotic | 4,400 (79.5) | 3,727 (78.8) | 1,986 (71.2) | 1,863 (70.7) | 673 (47.1) | 481 (44.9) | 6 (46.2) |
| Compensated cirrhosis | 472 (8.5) | 412 (8.7) | 307 (11.0) | 293 (11.1) | 477 (33.3) | 385 (35.8) | 1-5 (≤38.5) |
| Decompensated cirrhosis | 400 (7.2) | 362 (7.6) | 290 (10.4) | 281 (10.7) | 172 (12.0) | 125 (11.6) | 1-5 (≤38.5) |
| Hepatocellular carcinoma | 265 (4.8) | 232 (4.9) | 206 (7.4) | 199 (7.5) | 109 (7.6) | 83 (7.7) | 1-5 (≤38.5) |
| **HCV genotype, N (%)** |  |  |  |  |  |  |  |
| Genotype 1 | 1,431 (56.1) | 1,431 (56.1) | 1,431 (56.1) | 1,431 (56.1) | 762 (57.7) | 604 (60) | 1-5 (≤38.5) |
| Genotype 2 | 542 (21.2) | 542 (21.2) | 542 (21.2) | 542 (21.2) | 270 (20.5) | 202 (20.1) | 1-5 (≤38.5) |
| Genotype 3 | 239 (9.4) | 239 (9.4) | 239 (9.4) | 239 (9.4) | 121 (9.2) | 79 (7.8) | 1-5 (≤38.5) |
| Genotype 4 | 219 (8.6) | 219 (8.6) | 219 (8.6) | 219 (8.6) | 110 (7.7) | 81 (7.6) | 0 (.) |
| Genotype 5 | 21 (0.8) | 21 (0.8) | 21 (0.8) | 21 (0.8) | 9 (0.6) | 6 (0.6) | 0 (.) |
| Genotype 6 | 48 (1.9) | 48 (1.9) | 48 (1.9) | 48 (1.9) | 24 (1.7) | 17 (1.6) | 1-5 (≤38.5) |
| Coinfected | 52 (2.0) | 52 (2.0) | 52 (2.0) | 52 (2.0) | 24 (1.7) | 18 (1.7) | 0 (.) |
| **HCV diagnosis (first time), N (%)** |  |  |  |  |  |  |  |
| Pre-DAA era (<2014) | 4,304 (77.7) | 3,617 (76.4) | 2,419 (86.7) | 2,291 (86.9) | 1,243 (86.9) | 937 (87.2) | ≥8 (≥61.5) |
| DAA era (≥2014) | 1,233 (22.3) | 1,116 (23.6) | 370 (13.3) | 345 (13.1) | 188 (13.1) | 137 (12.8) | 1-5 (≤38.5) |
| **Treatment initiation (first time), N (%)** |  |  |  |  |  |  |  |
| Pre-DAA era (<2014) | 621 (43.4) | 621 (43.4) | 621 (43.4) | 601 (42.8) | 621 (43.4) | 425 (39.6) | ≥8 (≥61.5) |
| DAA era (≥2014) | 810 (56.6) | 810 (56.6) | 810 (56.6) | 803 (57.2) | 810 (56.6) | 649 (60.4) | 1-5 (≤38.5) |
| **Drug plan, N (%)** |  |  |  |  |  |  |  |
| Low income (Trillium or Special Drugs Program) | 126 (8.8) | 126 (8.8) | 126 (8.8) | 126 (8.8) | 126 (8.8) | 81 (7.5) | 0 (.) |
| Low income (Ontario Works) | 18 (1.3) | 18 (1.3) | 18 (1.3) | 18 (1.3) | 18 (1.3) | 9 (0.8) | 1-5 (≤38.5) |
| Senior (Senior and Low-Income Senior) | 993 (69.4) | 993 (69.4) | 993 (69.4) | 993 (69.4) | 993 (69.4) | 719 (66.9) | ≥8 (≥61.5) |
| Long-term care or home care | 14 (1.0) | 14 (1.0) | 14 (1.0) | 14 (1.0) | 14 (1.0) | 1-5 (≤0.5) | 0 (.) |
| OHIP+ (for 24 years and younger) | 0 (.) | 0 (.) | 0 (.) | 0 (.) | 0 (.) | 0 (.) | 0 (.) |
| Other public plan | 49 (3.4) | 49 (3.4) | 49 (3.4) | 49 (3.4) | 49 (3.4) | 29 (2.7) | 0 (.) |
| Unknown (Presumed private plan) | 231 (16.1) | 231 (16.1) | 231 (16.1) | 231 (16.1) | 231 (16.1) | 231 (21.5) | 0 (.) |

**S.Table 4b.**
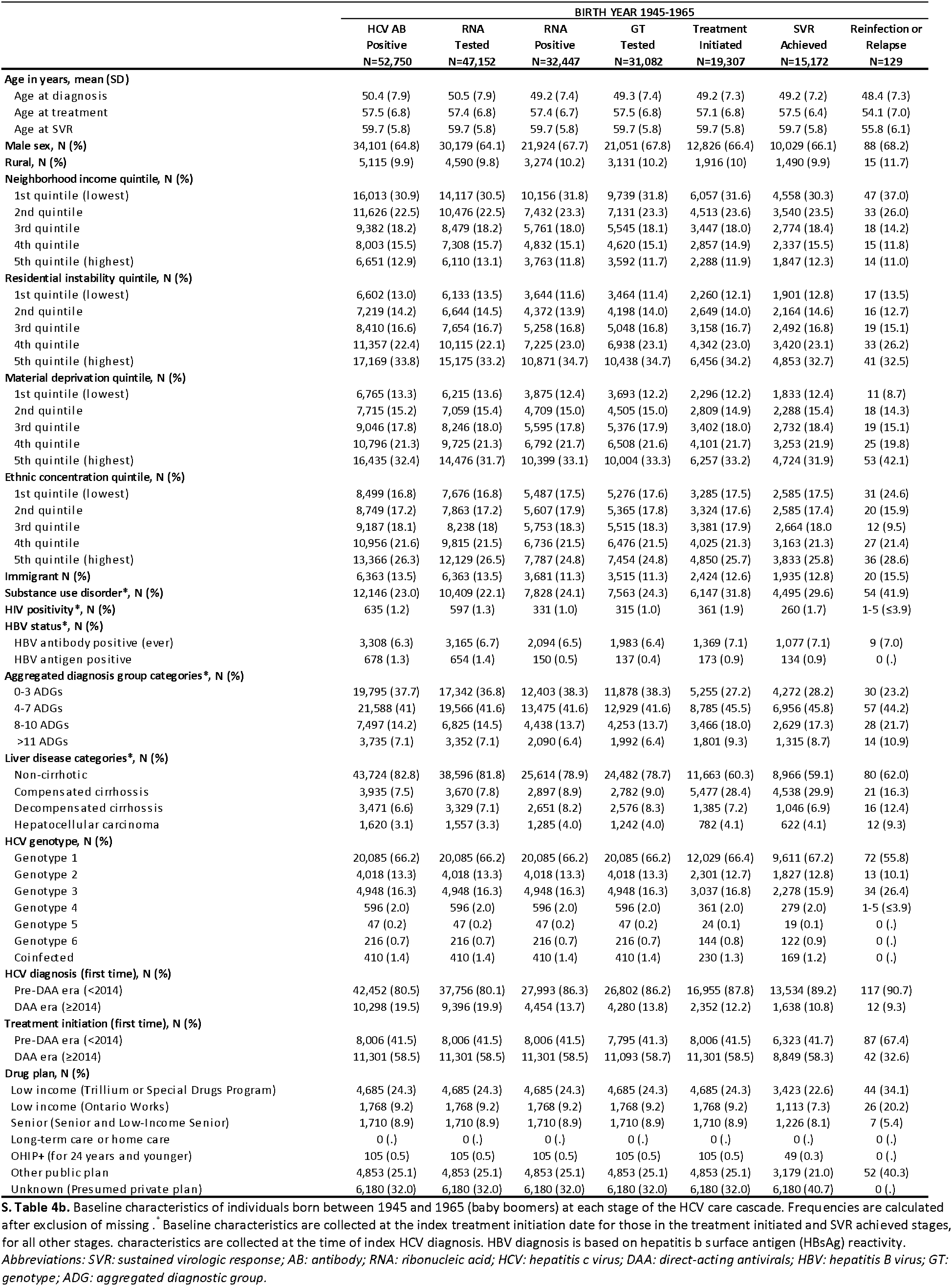
Baseline characteristics stratified by birth year (birth year 1945-1965)

**S.Table 4c.**
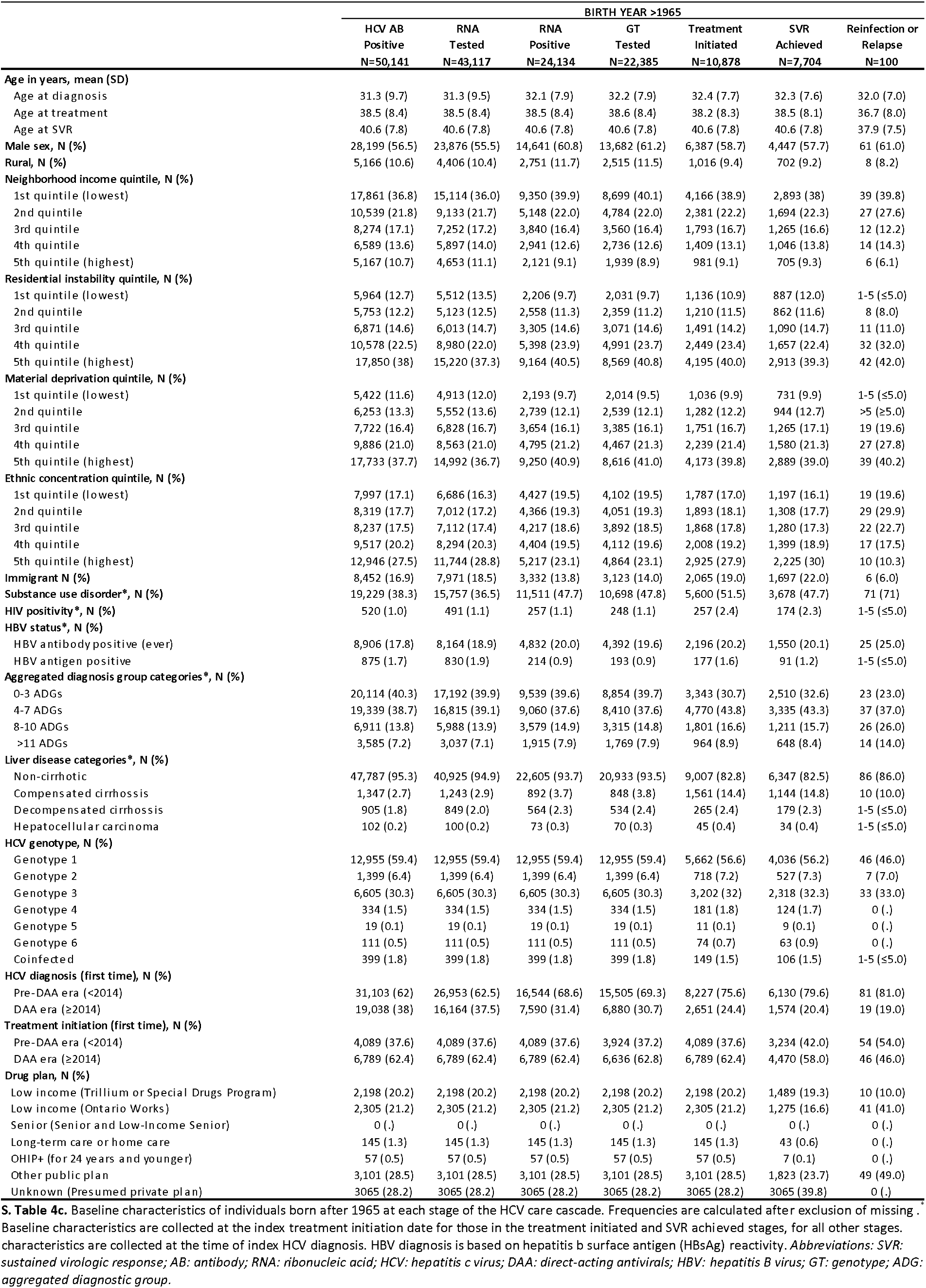
Baseline characteristics stratified by birth year (birth year > 1965)

**S.Table 5a.**
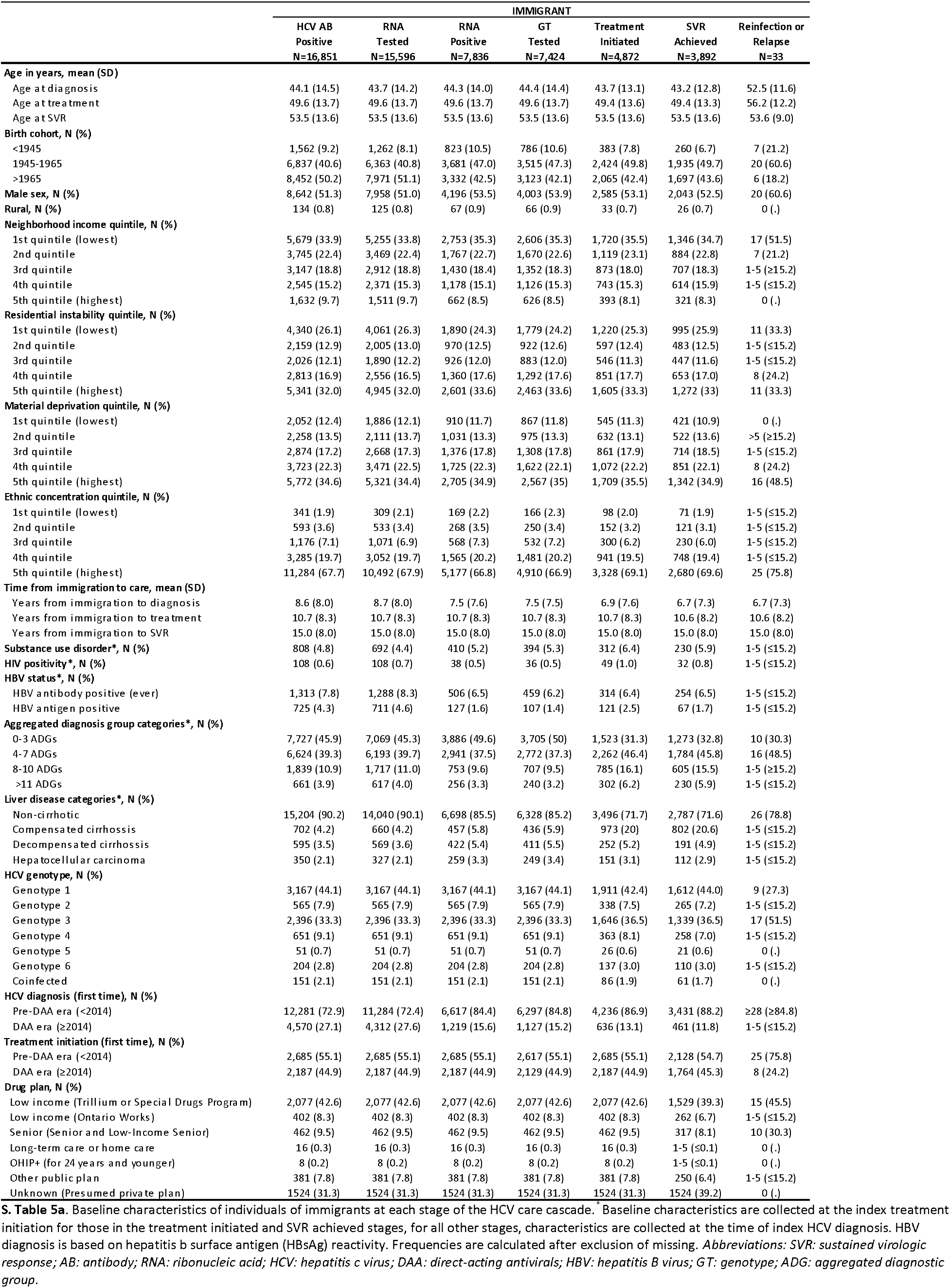
Baseline characteristics stratified by immigration status (immigrants)

**S.Table 5b.**
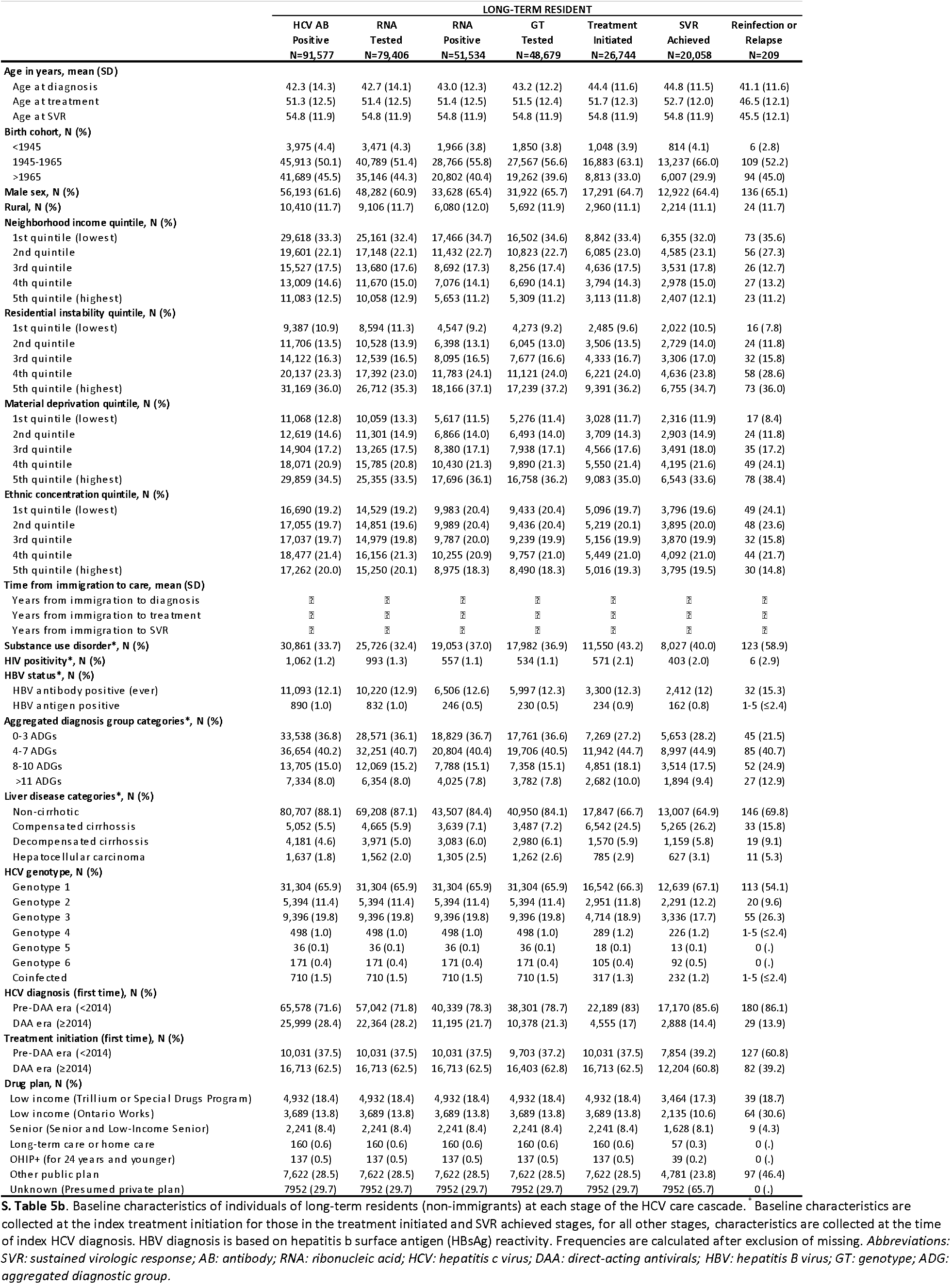
Baseline characteristics stratified by immigration status (long-term residents)

**S.Table 6a.** Baseline characteristics stratified by residential instability level (low instability)

|  | LOW RESIDENTIAL INSTABILITY |  |  |  |  |  |  |
| --- | --- | --- | --- | --- | --- | --- | --- |
|  | HCV AB | RNA | RNA | GT | Treatment | SVR | Reinfection or |
|  | Positive<br>N=27,592 | Tested<br>N=25,188 | Positive<br>N=13,805 | Tested<br>N=13,019 | Initiated<br>N=7,808 | Achieved<br>N=4,948 | Relapse<br>N=53 |
| **Age in years, mean (SD)** |  |  |  |  |  |  |  |
| Age at diagnosis | 44.2 (15.2) | 44.3 (15.0) | 45.0 (13.3) | 45.2 (13.1) | 45.7 (12.6) | 45.9 (12.2) | 48.1 (13.6) |
| Age at treatment | 52.4 (13.3) | 52.4 (13.3) | 52.5 (13.3) | 52.5 (13.3) | 52.7 (13.1) | 53.4 (12.6) | 52.9 (13.8) |
| Age at SVR | 56.0 (13.2) | 56.0 (13.2) | 56.0 (13.2) | 56.0 (13.2) | 56.0 (13.2) | 56.0 (13.2) | 54.8 (13.4) |
| **Birth cohort, N (%)** |  |  |  |  |  |  |  |
| <1945 | 2,054 (7.4) | 1,776 (7.1) | 1,025 (7.4) | 967 (7.4) | 553 (7.1) | 415 (6.6) | 8 (15.1) |
| 1945-1965 | 13,821 (50.1) | 12,777 (50.7) | 8,016 (58.1) | 7,662 (58.9) | 4,909 (62.9) | 4,065 (65.3) | 33 (62.3) |
| >1965 | 11,717 (42.5) | 10,635 (42.2) | 4,764 (34.5) | 4,390 (33.7) | 2,346 (30.0) | 1,749 (28.1) | 12 (22.6) |
| **Male sex, N (%)** | 15,941 (57.8) | 14,424 (57.3) | 8,638 (62.6) | 8,207 (63) | 4,828 (61.8) | 3,850 (61.8) | 28 (52.8) |
| **Rural, N (%)** | 3,151 (11.4) | 2,781 (11.0) | 1,841 (13.3) | 1,731 (13.3) | 956 (12.2) | 729 (11.7) | 8 (15.1) |
| **Neighborhood income quintile, N (%)** |  |  |  |  |  |  |  |
| 1st quintile (lowest) | 1,554 (5.7) | 1,414 (5.6) | 841 (6.0) | 790 (6.1) | 477 (6.2) | 370 (5.9) | 1-5 (≤9.4) |
| 2nd quintile | 4,241 (15.4) | 3,867 (15.4) | 2,191 (15.9) | 2,058 (15.8) | 1,313 (16.8) | 1,053 (16.9) | 15 (28.3) |
| 3rd quintile | 6,772 (24.5) | 6,161 (24.5) | 3,450 (25) | 3,269 (25.1) | 1,950 (25) | 1,573 (25.3) | ≥7 (≥13.3) |
| 4th quintile | 7,702 (27.9) | 7,036 (27.9) | 3,847 (27.9) | 3,634 (27.9) | 2,150 (27.5) | 1,721 (27.6) | 13 (24.5) |
| 5th quintile (highest) | 7,318 (26.5) | 6,705 (26.6) | 3,471 (25.2) | 3,264 (25.1) | 1,916 (24.5) | 1,511 (24.3) | 13 (24.5) |
| **Residential instability quintile, N (%)** |  |  |  |  |  |  |  |
| 1st quintile (lowest) | 13,727 (49.7) | 12,655 (50.2) | 6,437 (46.6) | 6,052 (46.5) | 3,705 (47.5) | 3,017 (48.4) | 27 (50.9) |
| 2nd quintile | 13,865 (50.3) | 12,533 (49.8) | 7,368 (53.4) | 6,967 (53.5) | 4,103 (52.5) | 3,212 (51.6) | 26 (49.1) |
| 3rd quintile | █ | █ | █ | █ | █ | █ | █ |
| 4th quintile | █ | █ | █ | █ | █ | █ | █ |
| 5th quintile (highest) | █ | █ | █ | █ | █ | █ | █ |
| **Material deprivation quintile, N (%)** |  |  |  |  |  |  |  |
| 1st quintile (lowest) | 6,737 (24.4) | 6,194 (24.5) | 3,266 (23.6) | 3,065 (23.5) | 1,810 (23.2) | 1,422 (22.8) | 9 (17.0) |
| 2nd quintile | 7,168 (26.0) | 6,540 (26.0) | 3,668 (26.6) | 3,473 (26.7) | 2,086 (26.7) | 1,690 (27.1) | 18 (34.0) |
| 3rd quintile | 6,363 (23.1) | 5,806 (23.1) | 3,261 (23.6) | 3,092 (23.7) | 1,877 (24.0) | 1,508 (24.2) | 12 (22.6) |
| 4th quintile | 4,888 (17.7) | 4,430 (17.6) | 2,385 (17.3) | 2,246 (17.3) | 1,340 (17.2) | 1,069 (17.2) | 8 (15.1) |
| 5th quintile (highest) | 2,436 (8.8) | 2,218 (8.8) | 1,225 (8.9) | 1,143 (8.8) | 695 (8.9) | 540 (8.7) | 6 (11.3) |
| **Ethnic concentration quintile, N (%)** |  |  |  |  |  |  |  |
| 1st quintile (lowest) | 3,868 (14.1) | 3,418 (13.6) | 2,274 (16.5) | 2,143 (16.4) | 1,194 (15.3) | 898 (14.3) | 10 (18.9) |
| 2nd quintile | 4,455 (16.1) | 3,975 (15.8) | 2,505 (18.1) | 2,369 (18.2) | 1,337 (17.1) | 1,056 (17.0) | 13 (24.5) |
| 3rd quintile | 4,582 (16.6) | 4,192 (16.6) | 2,417 (17.5) | 2,295 (17.6) | 1,327 (17.0) | 1,065 (17.1) | 1-5 (≤9.4) |
| 4th quintile | 5,557 (20.1) | 5,118 (20.3) | 2,660 (19.3) | 2,508 (19.3) | 1,521 (19.5) | 1,213 (19.5) | >5 (≥9.4) |
| 5th quintile (highest) | 9,130 (33.1) | 8,485 (33.7) | 3,949 (28.6) | 3,704 (28.5) | 2,429 (31.1) | 1,997 (32.1) | 18 (34.0) |
| **Immigrant N (%)** | 6,499 (23.6) | 6,066 (24.1) | 2,860 (20.7) | 2,701 (20.7) | 1,817 (23.3) | 1,478 (23.7) | 13 (24.5) |
| **Substance use disorder\*, N (%)** | 4,902 (17.8) | 4,185 (16.6) | 2,941 (21.3) | 2,776 (21.3) | 1,958 (25.1) | 1,416 (22.7) | 16 (30.2) |
| **HIV positivity\*, N (%)** | 152 (0.6) | 146 (0.6) | 66 (0.5) | 59 (0.5) | 80 (1.0) | 62 (1.0) | 1-5 (≤9.4) |
| **HBV status\*, N (%)** |  |  |  |  |  |  |  |
| HBV antibody positive (ever) | 2,731 (9.9) | 2,590 (10.3) | 1,286 (9.3) | 1,177 (9.0) | 706 (9) | 534 (8.6) | 10 (18.9) |
| HBV antigen positive | 599 (2.2) | 587 (2.3) | 99 (0.7) | 88 (0.7) | 97 (1.2) | 56 (0.9) | 0 (.) |
| **Aggregated diagnosis group categories\*, N (%)** |  |  |  |  |  |  |  |
| 0-3 ADGs | 10,453 (37.9) | 9,439 (37.5) | 5,401 (39.0) | 5,098 (39.1) | 2,147 (27.6) | 1,783 (28.6) | 10 (18.9) |
| 4-7 ADGs | 11,612 (42.1) | 10,691 (42.4) | 5,767 (41.8) | 5,440 (41.8) | 3,673 (47) | 2,934 (47.1) | 25 (47.2) |
| 8-10 ADGs | 3,787 (13.7) | 3,492 (13.9) | 1,843 (13.4) | 1,739 (13.4) | 1,353 (17.3) | 1,045 (16.8) | 13 (24.5) |
| >11 ADGs | 1,740 (6.3) | 1,566 (6.2) | 794 (5.8) | 742 (5.7) | 635 (8.1) | 467 (7.5) | 1-5 (≤9.4) |
| **Liver disease categories\*, N (%)** |  |  |  |  |  |  |  |
| Non-cirrhotic | 24,065 (87.3) | 21,828 (86.7) | 11,326 (82.1) | 10,654 (81.8) | 5,080 (65.1) | 3,944 (63.3) | 37 (69.8) |
| Compensated cirrhosis | 1,585 (5.7) | 1,487 (5.9) | 1,106 (8.0) | 1,043 (8.0) | 2,006 (25.7) | 1,726 (27.7) | 10 (18.9) |
| Decompensated cirrhosis | 1,332 (4.8) | 1,289 (5.1) | 926 (6.7) | 896 (6.9) | 447 (5.7) | 337 (5.4) | 1-5 (≤9.4) |
| Hepatocellular carcinoma | 610 (2.2) | 584 (2.3) | 447 (3.2) | 426 (3.3) | 275 (3.5) | 222 (3.6) | 1-5 (≤9.4) |
| **HCV genotype, N (%)** |  |  |  |  |  |  |  |
| Genotype 1 | 7,691 (60.7) | 7,691 (60.7) | 7,691 (60.7) | 7,691 (60.7) | 4,428 (61.1) | 3,619 (61.8) | 24 (45.3) |
| Genotype 2 | 1,500 (11.8) | 1,500 (11.8) | 1,500 (11.8) | 1,500 (11.8) | 799 (11.0) | 642 (11.0) | 1-5 (9.4) |
| Genotype 3 | 2,777 (21.9) | 2,777 (21.9) | 2,777 (21.9) | 2,777 (21.9) | 1,628 (22.5) | 1,280 (21.9) | 17 (32.1) |
| Genotype 4 | 355 (2.8) | 355 (2.8) | 355 (2.8) | 355 (2.8) | 200 (2.8) | 152 (2.6) | 1-5 (≤9.4) |
| Genotype 5 | 21 (0.2) | 21 (0.2) | 21 (0.2) | 21 (0.2) | 8 (0.1) | 8 (0.1) | 0 (.) |
| Genotype 6 | 149 (1.2) | 149 (1.2) | 149 (1.2) | 149 (1.2) | 95 (1.3) | 84 (1.4) | 0 (.) |
| Coinfected | 179 (1.4) | 179 (1.4) | 179 (1.4) | 179 (1.4) | 89 (1.2) | 67 (1.1) | 0 (.) |
| **HCV diagnosis (first time), N (%)** |  |  |  |  |  |  |  |
| Pre-DAA era (<2014) | 19,734 (71.5) | 17,999 (71.5) | 11,213 (81.2) | 10,616 (81.5) | 6,620 (84.8) | 5,428 (87.1) | 46 (86.8) |
| DAA era (≥2014) | 7,858 (28.5) | 7,189 (28.5) | 2,592 (18.8) | 2,403 (18.5) | 1,188 (15.2) | 801 (12.9) | 7 (13.2) |
| **Treatment initiation (first time), N (%)** |  |  |  |  |  |  |  |
| Pre-DAA era (<2014) | 3,406 (43.6) | 3,406 (43.6) | 3,406 (43.6) | 3,298 (43.4) | 3,406 (43.6) | 2,785 (44.7) | 31 (58.5) |
| DAA era (≥2014) | 4,402 (56.4) | 4,402 (56.4) | 4,402 (56.4) | 4,296 (56.6) | 4,402 (56.4) | 3,444 (55.3) | 22 (41.5) |
| **Drug plan, N (%)** |  |  |  |  |  |  |  |
| Low income (Trillium or Special Drugs Program) | 2,231 (28.6) | 2,231 (28.6) | 2,231 (28.6) | 2,231 (28.6) | 2,231 (28.6) | 1,638 (26.3) | 19 (35.8) |
| Low income (Ontario Works) | 635 (8.1) | 635 (8.1) | 635 (8.1) | 635 (8.1) | 635 (8.1) | 359 (5.8) | 9 (17.0) |
| Senior (Senior and Low-Income Senior) | 888 (11.4) | 888 (11.4) | 888 (11.4) | 888 (11.4) | 888 (10.5) | 635 (10.2) | 11 (20.8) |
| Long-term care or home care | 53 (0.7) | 53 (0.7) | 53 (0.7) | 53 (0.7) | 53 (0.6) | 16 (0.3) | 0 (.) |
| OHIP+ (for 24 years and younger) | 46 (0.6) | 46 (0.6) | 46 (0.6) | 46 (0.6) | 46 (0.5) | 15 (0.2) | 0 (.) |
| Other public plan | 1,094 (14.0) | 1,094 (14.0) | 1,094 (14.0) | 1,094 (14.0) | 1,094 (14.0) | 706 (11.3) | 14 (26.4) |
| Unknown (Presumed private plan) | 2860 (36.6) | 2860 (36.6) | 2860 (36.6) | 2860 (36.6) | 2860 (33.9) | 2860 (45.9) | 0 (.) |

**S.Table 6b.**
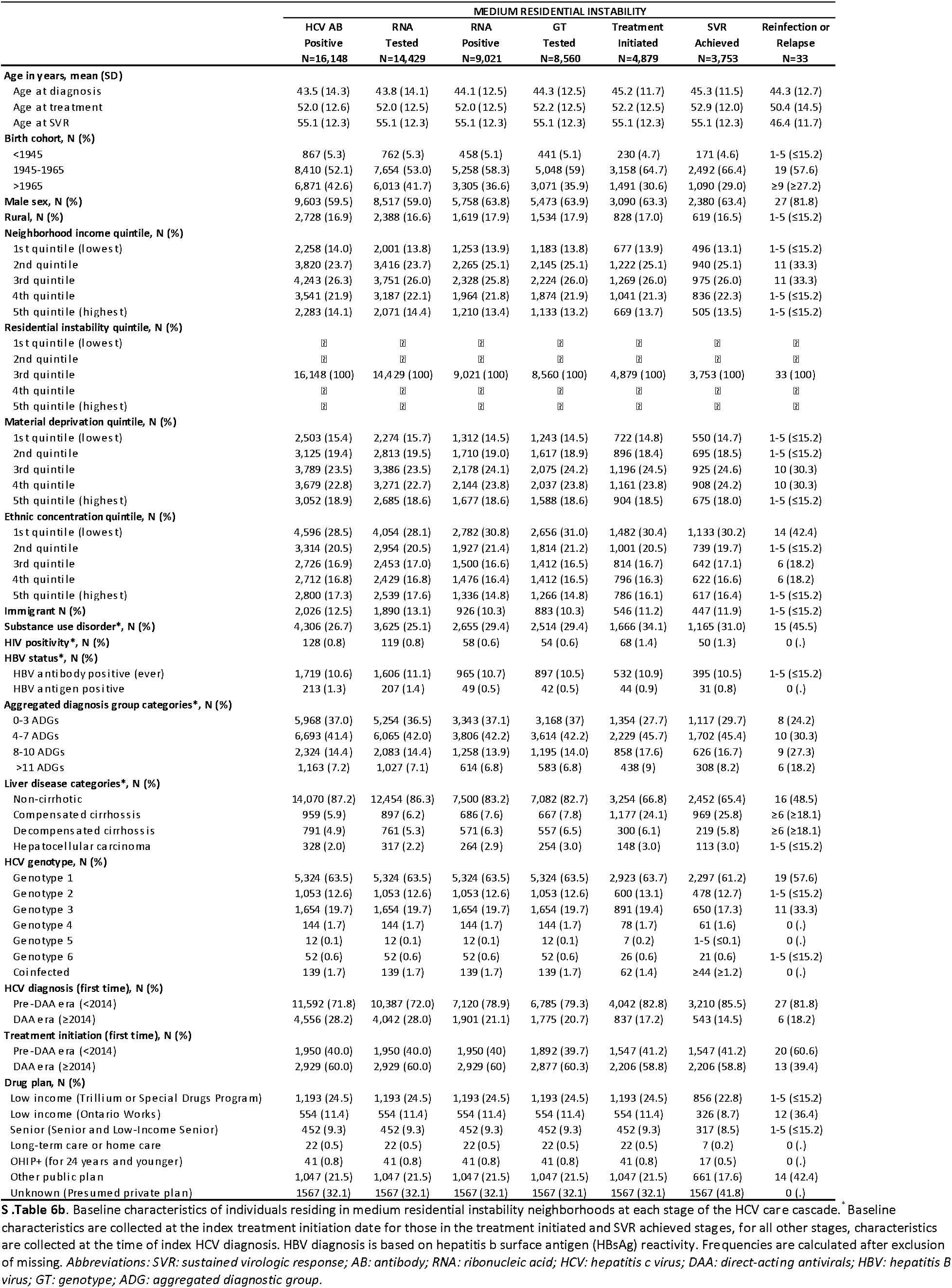
Baseline characteristics stratified by residential instability level (medium instability)

**S.Table 6c.**
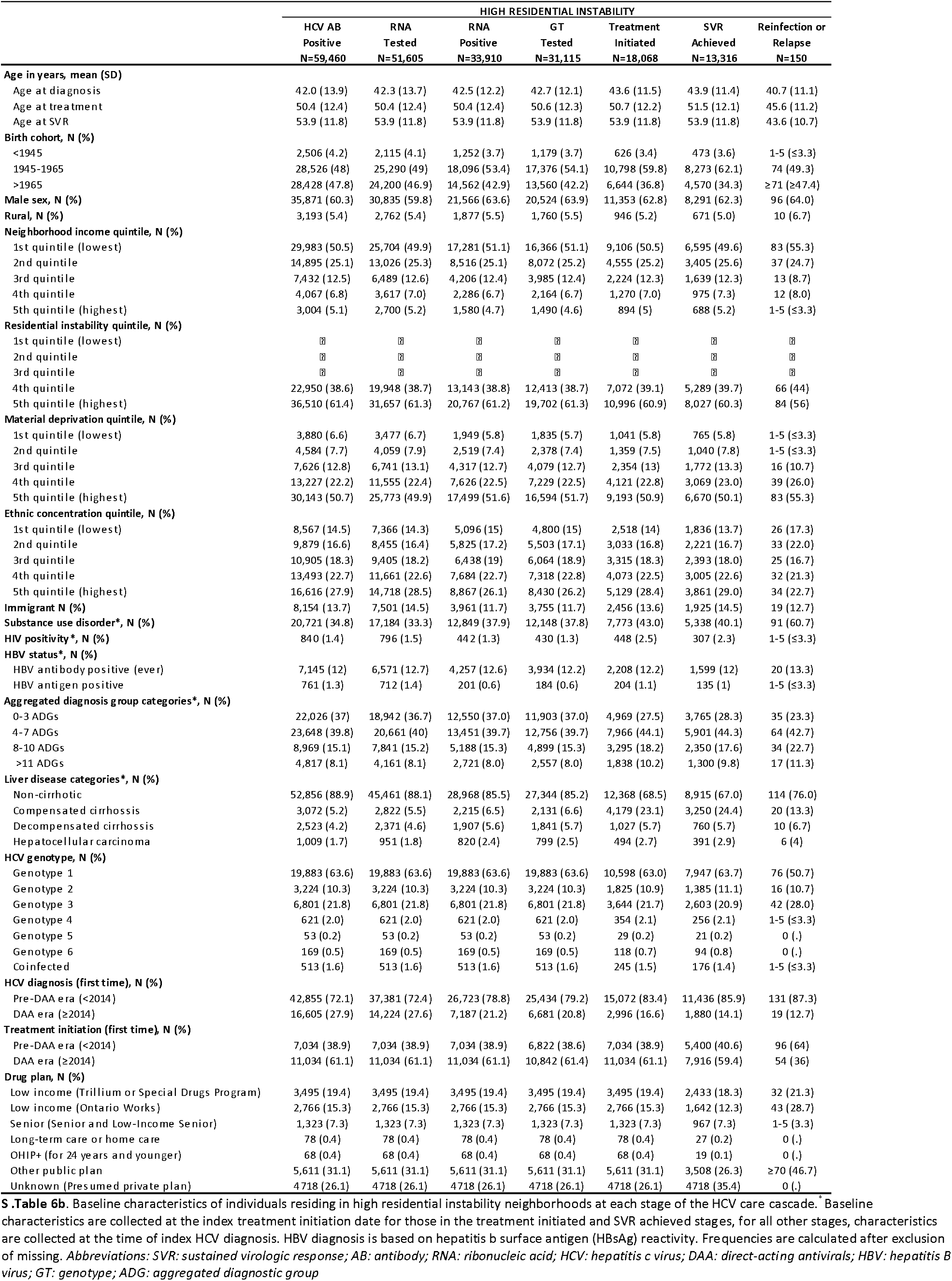
Baseline characteristics stratified by residential instability level (high instability)

**Supplemental Table 7.** Predictors of HCV antibody and RNA test positivity stratified by testing era.

| Covariates | AB Positivity |  | RNA Positivity |  |
| --- | --- | --- | --- | --- |
|  | Adjusted OR (95% CI) |  | Adjusted OR (95% CI) |  |
|  | Pre-DAA<br>N=841,548 | DAA<br>N=710,633 | Pre-DAA<br>N=81,746 | DAA<br>N=27,206 |
| **Age (yrs.)** |  |  |  |  |
| At time of diagnosis | **0.98 (0.98-0.99)** | **1.01 (1.01-1.01)** | 0.99 (0.98-0.99) | **1.01 (1.00-1.01)** |
| **Birth year** |  |  |  |  |
| 1945-1965 [ref.] | ? | ? | ? | ? |
| <1945 | **0.45 (0.43-0.46)** | **0.38 (0.36-0.41)** | **1.19 (1.10-1.28)** | **0.67 (0.59-0.77)** |
| >1965 | **0.20 (0.19-0.20)** | **0.77 (0.73-0.81)** | **0.44 (0.42-0.46)** | 0.98 (0.88-1.08) |
| **Sex** |  |  |  |  |
| Male [ref.] | ? | ? | ? | ? |
| Female | **0.67 (0.66-0.68)** | **0.71 (0.69-0.73)** | **0.68 (0.66-0.70)** | **0.71 (0.68-0.75)** |
| **Rurality** |  |  |  |  |
| Urban [ref.] | ? | ? | ? | ? |
| Rural | **0.58 (0.56-0.59)** | **0.83 (0.79-0.87)** | 1.02 (0.96-1.08) | 1.01 (0.92-1.11) |
| **Neighborhood income quintile** |  |  |  |  |
| Low (q. 1-2) [ref.] | ? | ? | ? | ? |
| Medium (q. 3) | **0.84 (0.82-0.6)** | **0.87 (0.81-0.87)** | **0.89 (0.86-0.94)** | **0.90 (0.82-0.98)** |
| High (q. 4-5) | **0.68 (0.66-0.7)** | 0.97 (0.93-1.01) | **0.75 (0.72-0.79)** | **0.84 (0.75-0.93)** |
| **Residential instability quintile** |  |  |  |  |
| Low (q. 1-2) [ref.] | ? | ? | ? | ? |
| Medium (q. 3) | **1.05 (1.03-1.08)** | **1.17 (1.12-1.22)** | **1.13 (1.07-1.18)** | **1.13 (1.04-1.21)** |
| High (q. 4-5) | **1.11 (1.11-1.14)** | **1.49 (1.42-1.55)** | **1.26 (1.21-1.31)** | **1.23 (1.04-1.21)** |
| **Material deprivation quintile** |  |  |  |  |
| Low (q. 1-2) [ref.] | ? | ? | ? | ? |
| Medium (q. 3) | **1.11 (1.09-1.14)** | **1.17 (1.12-1.22)** | 1.00 (0.95-1.05) | **1.15 (1.06-1.25)** |
| High (q. 4-5) | **1.22 (1.19-1.25)** | **1.49 (1.42-1.55)** | 1.00 (0.94-1.04) | **1.37 (1.25-1.49)** |
| **Ethnic concentration quintile** |  |  |  |  |
| Low (q. 1-2) [ref.] | ? | ? | ? | ? |
| Medium (q. 3) | 1.00 (0.98-1.02) | **0.90 (0.86-0.94)** | **0.91 (0.87-0.96)** | **0.93 (0.86-0.99)** |
| High (q. 4-5) | **1.06 (1.04-1.08)** | **0.75 (0.72-0.77)** | **0.77 (0.74-0.80)** | **0.67 (0.63-0.72)** |
| **Immigrant status** |  |  |  |  |
| Long-term resident [ref.] | ? | ? | ? | ? |
| Immigrant | 0.99 (0.97-1.01) | **0.84 (0.81-0.87)** | **0.86 (0.83-0.91)** | **0.72 (0.67-0.78)** |
| **Substance use disorder\*** |  |  |  |  |
| No [ref.] | ? | ? | ? | ? |
| Yes | **3.01 (2.96-3.07)** | **5.50 (5.34-5.66)** | **1.70 (1.63-1.77)** | **3.37 (3.17-3.59)** |
| **HIV positivity\*** |  |  |  |  |
| No [ref.] | ? | ? | ? | ? |
| Yes | **2.10 (1.93-2.28)** | **3.13 (2.60-3.77)** | **0.70 (0.62-0.80)** | **0.55 (0.43-0.78)** |
| **HBsAG positivity\*** |  |  |  |  |
| No [ref.] | ? | ? | ? | ? |
| Yes | **0.51 (0.48-0.56)** | **1.15 (1.02-1.31)** | **0.18 (0.16-0.21)** | **0.22 (0.18-0.28)** |
| **Total Number of ADGs (N.)\*** | **0.95 (0.95-0.95)** | **0.98 (0.98-0.99)** | **0.96 (0.96-0.96)** | **0.94 (0.93-0.95)** |
| **Liver disease\*** |  |  |  |  |
| No cirrhosis [ref.] | ? | ? | ? | ? |
| Compensated cirrhosis | **4.19 (4.04-4.35)** | **2.79 (2.61-2.99)** | **1.98 (1.84-2.13)** | **2.34 (2.06-2.66)** |
| Decompensated cirrhosis | **2.71 (2.62-2.79)** | **1.24 (1.15-1.33)** | **2.36 (2.21-2.52)** | 0.98 (0.88-1.11) |
| Hepatocellular carcinoma | **7.27 (6.90-7.65)** | **4.92 (4.41-5.49)** | **3.53 (3.14-3.96)** | **2.03 (1.69-2.45)** |

